# Multisensory Flicker Modulates Widespread Brain Networks and Reduces Interictal Epileptiform Discharges in Humans

**DOI:** 10.1101/2023.03.14.23286691

**Authors:** Lou T. Blanpain, Emily. Chen, James Park, Michael Y. Walelign, Robert E. Gross, Brian T. Cabaniss, Jon T. Willie, Annabelle C. Singer

## Abstract

Modulating brain oscillations has strong therapeutic potential. However, commonly used non-invasive interventions such as transcranial magnetic or direct current stimulation have limited effects on deeper cortical structures like the medial temporal lobe. Repetitive audio- visual stimulation, or sensory flicker, modulates such structures in mice but little is known about its effects in humans. Using high spatiotemporal resolution, we mapped and quantified the neurophysiological effects of sensory flicker in human subjects undergoing presurgical intracranial seizure monitoring. We found that flicker modulates both local field potential and single neurons in higher cognitive regions, including the medial temporal lobe and prefrontal cortex, and that local field potential modulation is likely mediated via resonance of involved circuits. We then assessed how flicker affects pathological neural activity, specifically interictal epileptiform discharges, a biomarker of epilepsy also implicated in Alzheimer’s and other diseases. In our patient population with focal seizure onsets, sensory flicker decreased the rate interictal epileptiform discharges. Our findings support the use of sensory flicker to modulate deeper cortical structures and mitigate pathological activity in humans.

## INTRODUCTION

The brain’s natural tendency to track dynamic sensory stimuli has untapped therapeutic potential. Rhythmic neural activity, or brain oscillations, play a key role in many brain functions including sensory processing^1^, attention, and memory^2^, and are impaired in diseases such as Alzheimer’s disease (AD), schizophrenia, and autism^3^. Driving or restoring brain oscillations associated with sensorimotor, mnemonic, and other cognitive processes has important therapeutic implications^4–20^. Because brain activity responds to sensory inputs, sensory stimulation has the potential to manipulate brain rhythms for therapeutic purposes. However, using sensory stimulation to drive brain rhythms in humans requires a clearer understanding of this naturally occurring process, including the extent of modulation across the brain and the mechanisms underlying such modulation, as well as the effects of sensory stimulation on pathological neural activity.

Audio-visual stimuli that flicker (turn on and off rhythmically) induce rhythmic neural activity, but it is unclear whether these sensory-induced oscillations occur within spatially constrained neural populations, or the extent of circuits affected. Steady-state evoked potential (steady-state EP), the periodic neurophysiological oscillation matching the on-off kinetics of a periodic stimulus^21^, has been implicated in processing of auditory stimuli such as speech^1^, and its abnormality is associated with neuropsychiatric conditions such as schizophrenia^22,23^ and autism^24^. This neurophysiological response has been extensively studied in humans albeit mostly with limited resolution in either spatial or temporal domains. Most prior studies of steady-state EP used scalp electroencephalography^25–35^ (EEG), where the steady-state EP results from summed activity of large, primarily sensory circuits. In the case of auditory stimulation for example, the steady-state EP could result from the staggered summation of signals spanning from brainstem to auditory cortex^32^. A limited number of studies have used intracranial EEG (iEEG) to spatially define responses to flicker in sensory circuits^36,37^, but few have examined the effects of sensory flicker in cognitive regions implicated in disease^38–40^.

There has been a long-standing debate about the mechanisms underlying the steady-state EP. One hypothesis posits that the steady-state EP emerges from the linear superposition of individual sensory single pulse evoked potentials (single pulse EPs)^41^. Another proposes that flicker entrains endogenous oscillations, implying flicker may affect functions associated with such oscillations^42^. Prior human studies have offered contradicting conclusions^40,43–45^, perhaps partly due to limited spatial resolution of the techniques employed. As a result, the mechanisms that underlie steady-state EP need to be clarified to understand both the neurophysiology of rhythmic audio-visual sensory processing and the potential of flicker to modulate brain functions for therapeutic applications.

The effects of sensory flicker on pathological neural activity are unclear. Of particular interest are interictal epileptiform discharges (IEDs) which are suspected to play a causal role in cognitive deficits^46,47^ and have increased prevalence in multiple brain diseases including epilepsy, AD^48,49^, autism, attention deficit hyperactivity disorder (ADHD), and multiple sclerosis^50^. Prior work provides conflicting insights into how flicker may affect IEDs. Restoring gamma oscillations in a mouse model of AD by rescuing the function of interneurons, reduced seizure activity and decreased memory deficits^4,5^. If driving gamma via sensory flicker has similar effects these prior results suggest that flicker could be used to reduce IEDs. Furthermore, in an AD mouse model, 40Hz sensory flicker was shown to modulate the medial temporal lobe (MTL) and prefrontal cortex (PFC), decrease the pathogenic peptide amyloid beta, and improve spatial memory deficits^51^. One study in humans suggested that visual flicker stimulation had an inhibitory effect on epileptic discharges in mesial temporal lobe epilepsy associated with hippocampal sclerosis^52^. Another small (4 subjects) pilot study^53^ suggested that exposure to 40Hz monotone decreases IEDs in epileptic patients. These studies raise the possibility that specific sensory stimuli could affect brain activity to potential therapeutic benefit. On the other hand, repetitive visual stimulation (particularly slower frequencies around 15Hz) may recruit and synchronize epileptogenic networks and trigger generalized seizures in genetically susceptible individuals. Prevalence of such susceptibility may be 1 in 10,000 individuals, or 2-14% in patients with known epilepsy^54^. However, few studies have robustly assessed whether different frequencies and modalities of flicker may decrease IEDs. For diseases like AD and epilepsy, where abnormal activity such as IEDs extends over multiple brain regions, sensory flicker may offer advantages over conventional brain stimulation approaches which are either invasive, limited to superficial brain structures, or limited in the extent of modulation. To determine how multisensory flicker of given modality-frequency combinations might impact normal and abnormal brain activity, a baseline understanding of mechanisms and impact upon distinct brain circuits is needed.

Accordingly, we examined the neurophysiological effects of visual and auditory flicker using stereoelectroencephalography (SEEG) in human subjects, the gold standard for recording neural activity with high spatial and temporal resolution. We recorded local field potentials (LFPs) across widespread cortical regions, and single neuron activity in the MTL and cingulate, while subjects were exposed to visual, auditory, and combined audio-visual flicker at multiple frequencies. We report that flicker affects neural activity across widespread brain structures, including those implicated in cognition, such as the MTL and PFC. Mechanistically, our findings are consistent with a model in which flicker-induced steady-state EP emerges from resonance of circuits rather than linear superposition of single pulse EPs. Finally, we found that flicker modestly but broadly decreases the overall rate of IEDs in epileptic patients. Thus, multisensory flicker modulates widespread brain networks and impacts pathological epileptiform neural activity in humans.

## RESULTS

### Audio-visual flicker modulates the human MTL and PFC

We investigated the effects of flicker on human brain activity by evaluating treatment-resistant patients with epilepsy undergoing pre-surgical intracranial seizure monitoring. In a first experiment, we exposed 12 participants to 10s trials of 5.5Hz, 40Hz, 80Hz, and random non-periodic visual (V), audio-visual (AV) and auditory (A) flicker, as well as baseline no stimulation (total of 13 conditions). During stimulation and baseline periods we recorded LFP from clinically targeted regions (1896 contact locations; Figure 1A, 1B, S1). Frequencies of 5.5Hz, 40Hz, and 80Hz were designed to mimic frequencies of endogenous theta, slow gamma, and fast gamma brain rhythms^55^, respectively, and random stimuli served as a non-periodic control. As a positive control, we first confirmed that flicker modulates canonical visual regions (including the pericalcarine, cuneus, lingual and lateral occipital cortices) and auditory regions (including transverse temporal or primary auditory cortex and the superior temporal cortex). 40Hz-V flicker modulated 58.1% of the contacts in visual areas, with median 5.1 fold-change in power at the frequency of stimulation relative to baseline (25^th^ and 75^th^ percentiles 1.5 and 19.2), compared to 13.7% of contacts in auditory areas with a median 0.6 fold-change in power (25^th^ and 75^th^ percentiles 0.4 and 5.5) (Figure 1C and 1D). Of note, some of the temporal modulation observed with 40Hz-V stimulation may be due to early visual processing from the third visual pathway in the superior temporal sulcus^56^. In contrast, 40Hz-A stimulation modulated 15.3% of contacts in auditory regions with median 0.9-fold increase in power (25^th^ and 75^th^ percentiles 0.6 and 4.6), compared to 3.2% of contacts in visual areas with median 0.4 fold-change (25^th^ and 75^th^ percentiles 0.3 and 0.5) (Figure 1E and 1F). These results confirmed that flicker stimulation effectively engages expected sensory regions.

**Figure 1.**
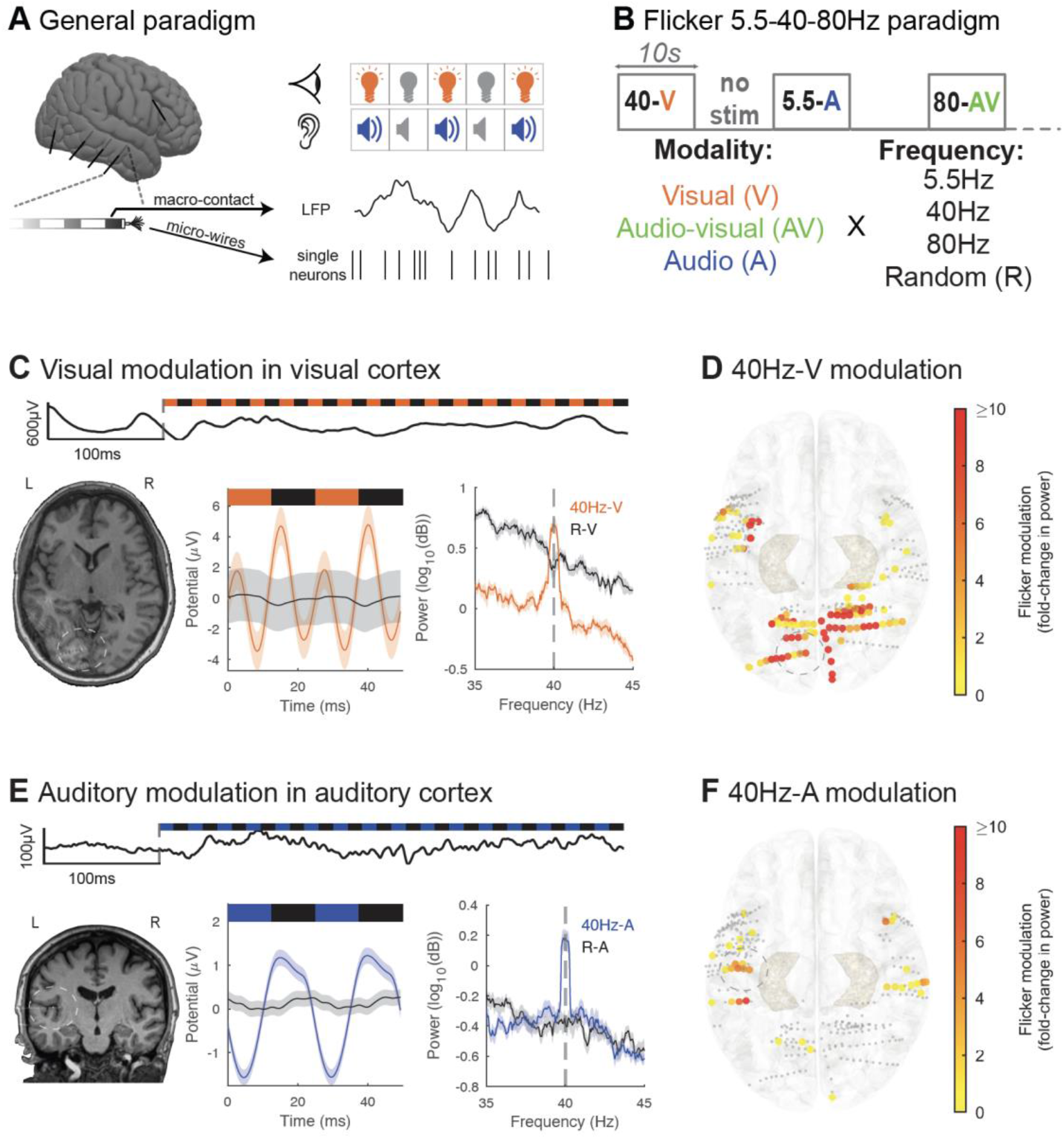
Audio-visual flicker induces steady-state evoked potential in human sensory regions. (A) Intracranial local field potential (LFP) and single neuron activity were recorded while we exposed subjects to visual and auditory stimulation pulses. (B) In this first paradigm, we exposed subjects to 10s trials of visual (V, orange), audio-visual (AV, green) or auditory (A, blue) flicker at 5.5Hz, 40Hz, 80Hz and random non-periodic stimuli, as well as no stimulation or baseline (total of 13 conditions). (C) Example of 40Hz-V steady-state evoked potential (EP) in early visual area lingual gyrus in one subject. Top: raw LFP trace at the beginning of a 40Hz-V trial. Bottom left: post-operative computed tomography (CT) scan overlaid on pre-operative magnetic resonance imaging (MRI), with contact from which results are shown highlighted with white circle. Bottom middle: LFP response to 40Hz-V flicker (orange) versus random visual stimulation (black), averaged over 2 cycles of the stimulus. Bottom right: average power spectral density (PSD) plot of 40Hz-V flicker versus random visual stimulation. For these last two plots, solid lines represent the mean and shaded areas represent standard error of the mean. (D) Response to 40Hz-V stimulation across contacts (represented by dots) located in early visual and auditory areas, represented on the Montreal Neurological Institute (MNI) normalized brain (top view), with color representing fold-change in power at the frequency of stimulation, capped at 10-fold increase to best visualize this range. Smaller grey dots indicate channels with no significant response. 33 contacts had modulation greater than 10-fold. The contact from which results are represented in (C) is highlighted with a grey circle. (E) Same as in (C) but illustrating 40Hz-A steady-state EP in Heschl’s gyrus or transverse temporal gyrus (primary auditory area) from one subject. Blue represents 40Hz auditory stimulation, while black represents random auditory stimulation. (F) Same as (D) but showing response to 40Hz-A stimulation. One contact had modulation greater than the 10-fold threshold. Contact from which results are represented in (E) is highlighted with a grey circle.

We then investigated whether sensory flicker modulates activity in higher cognitive regions frequently sampled in our subject population of focal-onset epilepsy patients, specifically the MTL and PFC. We found that flicker consistently alters LFP in these structures, with increased spectral power at the frequency of stimulation compared to baseline (Figure 2A and 2B). Across 294 MTL and 432 PFC contacts from 12 subjects, we found 40Hz-AV flicker significantly modulated 15.0% and 7.9% of contacts, respectively, with a median 1.1-fold increase (25^th^ and 75^th^ percentiles 0.5 and 1.7) and 0.4-fold increase (25^th^ and 75^th^ percentiles 0.3 and 0.8) in power relative to baseline, respectively for MTL and PFC (Figure 2C, 2D). These results were not explained by volume conduction of electrical potentials from nearby sensory-processing areas or by artifacts from our stimulation device, as we controlled for both by using off-line Laplacian re-referencing (see Methods) and by showing absence or reduction of a response under a relative occluded stimulation (Figure S1A). It is possible that some of the contacts lateral to the hippocampus may be picking up signal from optic radiations^57^, which are involved in early visual processing. Significant modulation was often highly localized to one or a few contacts on a recording probe (Figure 2A, 2B). As LFPs are considered to represent synchronized currents from organized dendritic inputs^58^, these results show that the MTL and PFC are modulated by sensory flicker despite not being commonly considered as directly involved in primary sensory processing.

**Figure 2.**
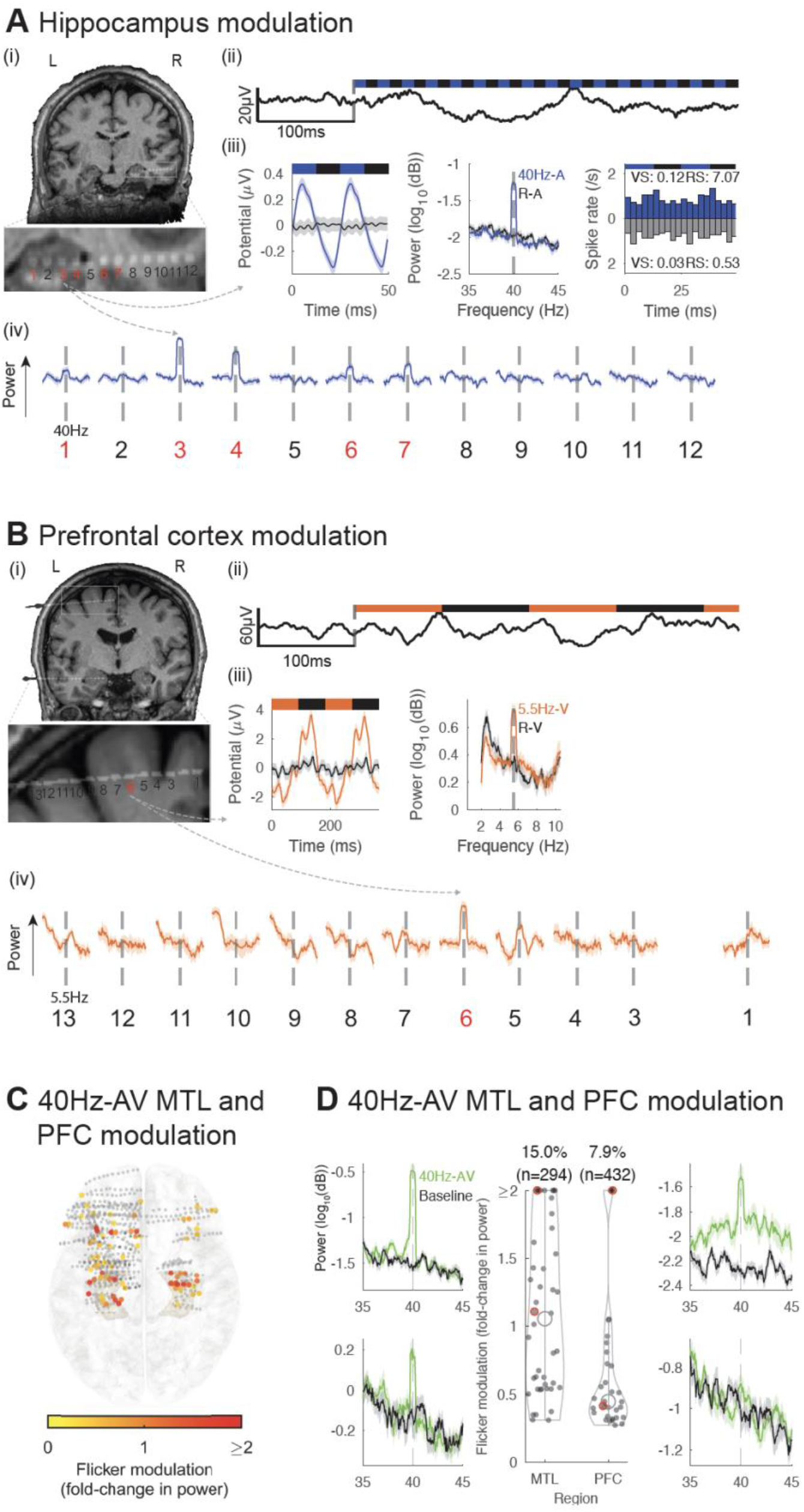
Audio-visual flicker induces steady-state evoked potential in the human medial temporal lobe and prefrontal cortex. (A) Example of 40Hz-auditory (A) steady-state evoked potential (EP) in the hippocampus (HPC). (i) location of a depth electrode with contacts numbered from deep to superficial (zoomed-in below), in an individual subject, overlaid on pre-operative magnetic resonance imaging (MRI); contacts 1 through 5 are in or near the HPC. (ii) example of a raw local field potential (LFP) trace for the beginning of a 40Hz-A trial. (iii) for the same contact, averaged evoked potential over 2 cycles of the stimulus (left), averaged power spectral density (PSD) during 40Hz-A flicker (blue) and random auditory (black, middle). Peristimulus time histogram (PSTH) for an example single neuron isolated in the hippocampus with corresponding vector strength (VS) and Rayleigh statistics (RS, right). (iv) averaged PSD for each contact from the depth electrode during 40Hz-A flicker, zoomed-in to show frequency of stimulation +/- 5Hz (solid line indicates mean and shaded area represents standard error of the mean), showing evoked responses in contacts 3, 4, in the HPC, and weaker response in contacts 1, 6 and 7 (marked in red). (B) Same as (A), but for a depth electrode in a different subject, targeting the superior prefrontal cortex (PFC), in response to 5.5Hz-visual (V) flicker (orange). (C) Across 12 subjects, electrode contacts (represented by dots) located in the medial temporal lobe (MTL) and prefrontal cortex (PFC) that were modulated by 40Hz-audiovisual (AV) flicker, represented on a 3D model of the Montreal Neurological Institute (MNI) normalized brain (top view), with color representing fold-change in power at the frequency of stimulation, capped at 2-fold increase to best visualize this range. Smaller grey dots indicate contacts with no significant response. (D) Middle: distribution of fold-change in power (capped at 2-fold change to best visualize this range) at the frequency of stimulation during 40Hz-AV flicker relative to baseline, for all electrode contacts shown in (C) that showed significant modulation. Percentages above violin plot indicate percent of electrodes showing significant steady-state EP in corresponding brain regions; black open circles represent medians of the distributions, filled dots indicate each contact with significant modulation. 8 contacts had modulation higher than capped 2-fold change in the MTL, and 4 contacts in the PFC. Left and right: example power spectral density plots of the response to 40Hz-AV flicker in the MTL and PFC, respectively; green and black represent 40Hz-AV condition and baseline, respectively. Solid lines represent the mean and shaded areas represent standard error of the mean. These examples are highlighted with red circles in the middle violin plots.

We evaluated whether spiking of single neurons in higher cognitive regions, specifically the hippocampus and cingulate, was modulated by flicker. While LFP primarily represents local summated dendritic inputs^58^, these inputs may or may not alter spiking output of individual neurons. To study neuron spiking activity, 4 subjects were also implanted with depth electrodes containing microwires (Figure 1A) that terminated in the hippocampus and cingulate, and a total of 25 units (13 single units, 12 multi-units) were isolated. Out of them, 21 units (7 single units and 3 multi-units in the hippocampus, 5 single units and 6 multi-units in the cingulate) had a spike rate that was high enough to assess modulation (see Methods). We found that some hippocampal and cingulate units showed higher firing probability at a given phase of the sensory stimulus for some frequencies and modalities of flicker compared to the random condition (Figure 2A, S2). These results suggest that flicker may modulate spiking activity of a subset of neurons located within higher cognitive regions in humans.

### Audio-visual flicker induces steady-state EPs across widespread functional networks

Next, we studied the extent of flicker modulation across the brain, and whether some regions were more responsive to specific modalities and frequencies of stimulation than others, which is important for understanding what regions can be targeted with this approach. Overall, audio- visual flicker produced the broadest responses across all frequencies tested, followed by visual flicker and auditory flicker (Figure 3A, top). With respect to frequencies of stimulation tested, more contacts exhibited a steady-state EP in response to 40Hz, than to 5.5Hz or 80Hz stimulation (Figure 3A, center). For locations responding to both visual and auditory flicker, the majority (63.1%) showed preferential responses to the same stimulation frequency (Figure 3A, bottom), suggesting that brain regions are sensitive to given frequencies of stimulation, irrespective of the modality. We determined the relative strength of steady-state EP to flicker by spatial distribution, modality, and frequency (Figure 3B). As expected, we observed a strong response in the occipital region for conditions involving the visual modality, as well as moderate responses of the parietal, temporal and prefrontal regions; the auditory-only condition mainly affected temporal and prefrontal regions; the 40Hz condition appeared to broadly impact most regions, particularly when using combined visual and auditory modalities. We also determined the strength 40Hz-AV flicker-induced steady-state EP by functional networks (Figure 3C) that were previously defined by fMRI resting state functional connectivity across 1,000 healthy subjects^59^. More than half (58.8%) of the contact locations in or near the visual network showed significant modulation to flicker with some regions having a more than 10-fold increase in power. More notably, flicker affected subsets of contact locations throughout networks not thought to be primarily involved in sensory processing, with 16.4%, 14.0%, 14.4%, 13.4.8% and 6.1% of the contacts showing significant steady-state EP in the ventral-attention, dorsal-attention, default, limbic, and frontoparietal networks, respectively.

**Figure 3.**
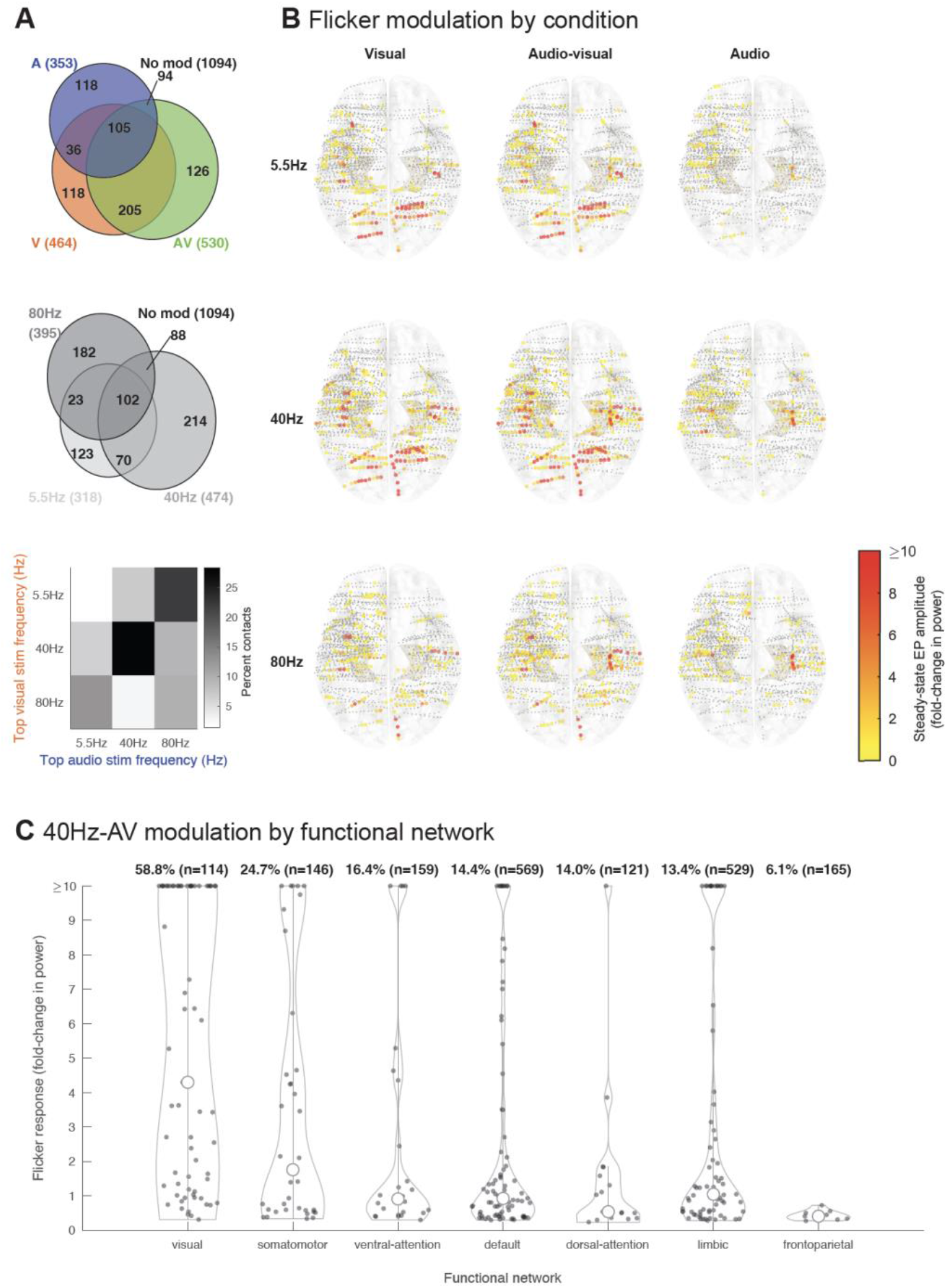
Flicker steady-state evoked potential across the brain. (A) Top: Venn diagram showing proportion of contacts (12 subjects, 1896 contacts) with significant steady-state evoked potential (EP) to visual (V, orange), audio-visual (AV, green) and audio (A, blue) flicker; absolute number of contacts are also shown. Center: Venn diagram showing significant responses to different flicker frequencies (5.5Hz-light grey, 40Hz-darker grey, 80Hz-dark grey). Bottom: comparison of top stimulation frequency for those contacts that responded to both visual and auditory flicker. Most contacts showed a preference for the same stimulation frequency when stimulated with either modality. (B) Approximate location of each contact (represented by dots) and associated amplitude of steady-state EP, plotted on Montreal Neurological Institute (MNI) normalized brain (top view), for each of 9 conditions: 5.5Hz, 40Hz, 80Hz stimulation frequencies at visual (V), audio-visual (AV) and auditory (A) modalities. Smaller grey dots indicate no significant response, while larger dots indicate significant steady-state EP, from yellow to red (0-10-fold or more increase in power). (C) Distribution of 40Hz-AV flicker steady-state EP across all contacts showing significant modulation from all subjects, categorized by functional networks (as previously defined by resting state functional connectivity characterized across 1,000 healthy subjects^59^). Percentages indicate percent of contacts in that network with significant responses, with absolute number of contacts localized to those networks in parentheses. Open circles represent medians of the distributions, filled dots indicate each significant contact.

### The steady-state EP does not result from linear superposition of single pulse EPs

Next, we tested potential mechanisms by which flicker induces a steady-state EP. One common hypothesis is that the steady-state EP results from the linear superposition of single pulse EPs^41,60,61^ (Figure 4A). This contrasts with another proposed mechanism, in which the steady-state EP results from intrinsic properties of circuits which have greater responses to specific stimulation frequencies. We first tested predictions of the linear superposition mechanism. Specifically, this mechanism predicts that 1) a region showing a single pulse EP should also show a steady-state EP, 2) the amplitude of the steady-state EP is proportional to that of the single pulse EP, meaning a region showing a strong sensory response to single pulses should show a correspondingly strong response to flicker, 3) the steady-state EP amplitude can be approximated by simulating the linear superposition of single pulse EPs, 4) the amplitude of the response to flicker decreases as the stimulation frequency increases, due to the low-pass filter properties of neurons and circuits^62^, and 5) there is no interaction between steady-state EP and endogenous oscillations, but rather a simple superposition or co-existence of the two.

**Figure 4.**
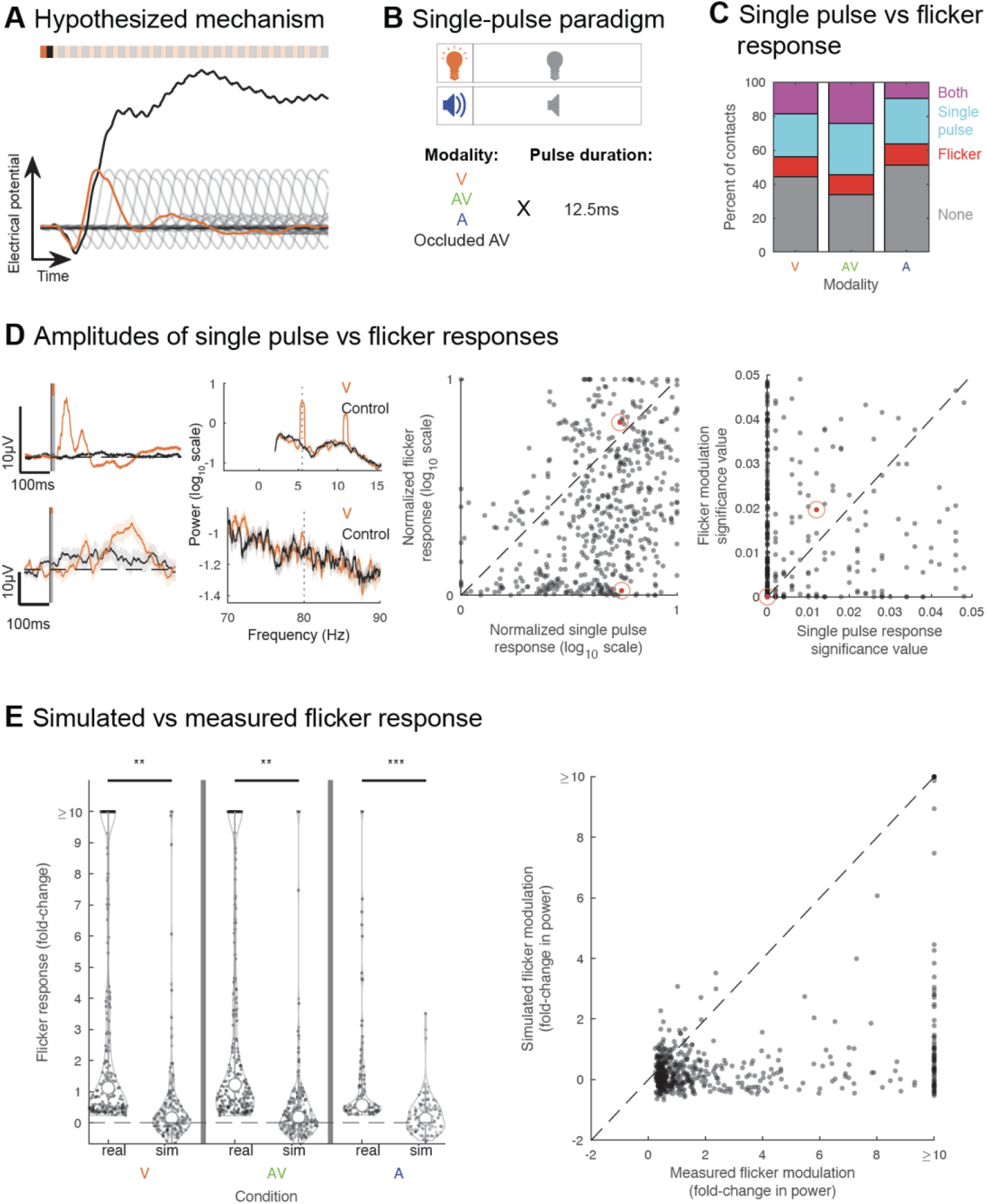
Flicker modulation does not result from linear superposition of single pulse evoked potentials. (A) We tested the hypothesis that the steady-state evoked potential (EP) is mediated by linear superposition of single pulse EPs: schematic of the response to a single visual pulse (orange), subsequent underlying single pulse responses if they repeated every 25ms (transparent grey), and the resulting response from linear summation of each pulse’s EP every 25ms, i.e., imitating 40Hz-visual flicker (black). (B) We ran an additional paradigm in a subset of 6 subjects, which involved exposure to single 12.5ms pulses in the visual, audio-visual, and auditory modalities; relative occluded exposure (subject wearing a sleep mask and earplugs) to audiovisual pulses was used as control. (C) Percent of contacts showing response to flicker-only (red), single pulse-only (cyan), both flicker and single pulses (purple), and no response (grey), with the visual, audiovisual, and auditory modality, respectively. (D) Left: Example of contacts that responded strongly to both (top) single visual pulses (left) and visual flicker (right) or more strongly to single pulses (bottom); stimulation condition in orange and control condition in black (relative occluded audio-visual (AV) for the single-pulse EP, random AV for power spectral density (PSD) plots). Top contact showed a strong response to visual stimulation, both in the case of single-pulses and flicker, while bottom contact showed a strong response to single-pulses but weak response to flicker. Solid line indicates mean and shaded area indicates standard error of the mean. Middle: scatter plot comparing the amplitude of steady-state EP versus amplitude of single-pulse EP, normalized by subject and stimulation modality, on a logarithmic scale; each dot represents a contact’s responses for a given modality; only contacts with both significant steady-state EP and single-pulse EP were included. Dots highlighted in red indicate the examples represented to the left. Right: scatter plot comparing, for the same contacts, significance values of flicker versus single pulse response. (E) Left: Distribution of amplitudes of flicker steady-state EP at 40Hz in the visual, audio-visual, and auditory modalities, using real data and simulated data across contacts from 6 subjects based on the linear superposition hypothesis. Only contacts showing significant flicker modulation in the real data were included; ** and *** correspond to p-value<0.01 and p-value<0.001, respectively; open circles represent medians of the distributions, dots indicate each contact. Right: for those same contacts, across modalities (each dot represents a contact’s real and simulated responses for a given modality), comparison between the amplitude of flicker steady-state EPs calculated using real data (x-axis) and using simulated data (y-axis).

To test the superposition hypothesis, we ran an additional experiment in a subset of 6 subjects (total of 1025 contact locations), where we exposed them to single pulses of visual, audio-visual, and auditory modalities (Figure 4B, S1B, S3A). We then quantified, for each contact, whether there was a single pulse EP and the magnitude of that response (see Methods). As expected, we found stronger responses in the occipital region with visual modality, and in the temporal region with auditory modality (Figure S3B). Surprisingly, we found widespread single-pulse EP beyond sensory regions. Moreover, we found that among locations showing any sensory response, 45.7-54.9% responded to single pulses only (45.7%, 46.1% and 54.9% for visual, audio- visual and audio modalities respectively), while 17.4-25.8% showed response to flicker only (21.1%, 17.4% and 25.8%) and 19.3-36.5% responded to both single pulse and flicker (33.2%, 36.5% and 19.3%) (Figure 4C, S3C). This indicates a discrepancy between single pulse versus flicker responses for most regions sampled, which is inconsistent with the superposition hypothesis, and may imply involvement of additional mechanisms, such as sensory adaptation^63^ or other modulatory dynamics specific to circuits involved in the processing of the stimulus.

We then considered if the superposition mechanism specifically explained responses of the subset of locations showing both a significant steady-state EP and single-pulse EP, rather than all recorded responses. If true, we would expect proportional amplitudes of the steady-state and single-pulse EPs in this subset of locations. We found, however, that the normalized log-scaled amplitudes of the flicker response and single-pulse EP were significantly different (paired-sample, two-sided t-test, p-value=0, confidence interval(ci)=[0.6270 0.7830], t-statistic (tstat)=17.7564, degrees of freedom (df)=530, standard deviation (sd)=0.9150), and that the significance of those responses also were different (paired-sample, two-sided t-test, p-value=6.3022×10^−8^, ci=[-0.0050-0.0024], tstat=-5.4882, df=530, sd=0.0155; Figure 4D). Most recorded locations showed a trend for a stronger single-pulse EP compared to steady-state EP, in contrast to the prediction of the superposition hypothesis.

One possible explanation for stronger single-pulse versus steady-state EP is destructive interference, where a peak of one pulse EP coincides with a trough of the previous pulse EP, resulting in an overall attenuated response in the case of steady-state EP. We tested, for each recording site showing significant 40Hz steady-state EP, whether we could artificially generate the expected flicker response based on the single pulse EP shape and amplitude for each of those contacts, as predicted by the superposition mechanism (see Methods). Such a simulation should account for destructive interference, and highlight whether this would explain low flicker amplitude response compared to single-pulse EP. We found that overall, out of recording sites showing significant 40Hz modulation, only a minority showed significant modulation in the simulated data (24.5%, 25.1% and 18.4% respectively in the visual, audio-visual and auditory conditions), and at a lower amplitude (0.2 versus 1.1, 0.2 versus 1.2, 0.3 versus 0.6 median fold-changes, respectively for visual, audio-visual and auditory conditions). The distribution of amplitudes of steady-state EP was significantly higher in the real data compared to the simulated data (Figure 4E, left; paired-sample, two-sided t-test of the real vs simulated data non-capped distributions for the visual condition p-value=0.0093, ci=[4.9421 34.7771], tstat=2.6246, df=207, sd=109.1273, for the audio-visual condition p-value=0.0081, ci=[4.9576 32.8400], tstat=2.6703, df=242, sd=110.3258, for the auditory condition p-value=2.5473×10^−6^, ci=[0.7363 1.7096], tstat=4.9847, df=102, sd=2.4900), and those values were significantly different from each other overall (Figure 4E, right; paired, two-sided t-test, p-value=1.6655 × 10^−4^, ci=[7.6969 24.2496], tstat=3.7910, df=553, sd=99.1734). This provides additional evidence against the linear superposition hypothesis and suggests that destructive interference does not explain the overall lower amplitude of the flicker response compared to single-pulse EP. Overall, our data does not fit the expected outcomes from the linear superstition hypothesis, making this an unlikely underlying mechanism for flicker modulation.

### Resonance of circuits to specific flicker frequencies

Since the steady-state EP was not explained by linear superposition of single pulse EPs, we evaluated if the steady-state EP could be explained by intrinsic oscillatory properties of circuits involved in processing of the stimulus. This hypothesis predicts that a single pulse EP is not required to observe a steady-state EP, and that the flicker response could show resonance^12,62^ or entrainment^1,16,43,64–66^, both related to intrinsic oscillatory circuit properties. We defined resonance as the preferential response of a circuit to specific frequency bands of stimulation resulting from the underlying cellular composition and their synaptic connections^62^. We predicted that resonance would manifest as 1) a stronger steady-state EP at a subset of flicker frequencies tested and 2) a stronger phase-locking value (PLV) between stimulus and LFP at similar frequencies. While the term entrainment can be used in a variety of ways, we defined entrainment of endogenous oscillations in the narrow sense^1^ as the ability of a repetitive stimulus to modulate an endogenous oscillation. We expected that entrainment would manifest as 1) a stronger change in power and 2) a stronger phase-locking with stimulation at a frequency close to that of an endogenous oscillation.

To test these predictions, we exposed a separate subset of 5 subjects (total of 821 contact locations across 6 sessions) to visual or auditory flicker at 26 different frequencies spanning the 5.5-80Hz range in about 3Hz intervals (Figure 5A and S1B), as well as random flicker and baseline (no stim), and estimated the amplitude of the steady-state EP resulting from each stimulation frequency. First, we showed that most recorded locations showing a steady-state EP have a preference to specific frequencies of stimulation (Figure 5B and 5C), in line with the resonance hypothesis and at odds with the superposition hypothesis. We found that most contacts across sessions show modulation, with 78.1% and 85.6% showing significant fold-change in power or PLV to the stimulus, respectively, for at least one stimulation frequency (Figure 5C). The stimulation frequencies with the strongest responses spanned the whole range tested from 5.5-80Hz. Moreover, 18.2% and 26.3% of contacts showed significant fold-change in power or PLV (median of 2.0 and 0.305, range of 0.3-545.3 and 0.075-0.970, 25^th^ percentile of 0.9 and 0.199, 75^th^ percentile of 5.7 and 0.510, respectively) to more than 6 of the stimulation frequencies tested. Of these only 3.2% and 5.2% showed a preference or strongest response to the lowest frequency of stimulation and otherwise most showed a preference for a variety of frequencies. Other contacts showed a preference for fewer (1-6) stimulation frequencies (59.9% and 59.3% of contacts showing significant fold-change or PLV, respectively), albeit their modulation values were lower (median of 0.5 and 0.135, range of 0.2-6.6 and 0.06-0.344, 25^th^ percentile of 0.4 and 0.116, 75^th^ percentile of 0.7 and 0.158, respectively). We also explored whether specific regions might respond preferentially to specific stimulation frequencies, among channels that showed significant fold-change at the frequency of stimulation, for more than six of the stimulation frequencies tested (Figure S4). We did not observe a clear clustering of stimulation frequency preference by brain region, regardless of the modality of stimulation (visual or auditory) used. Overall, most contact locations showed preferential response to stimulation at a given frequency, supporting a mechanism involving resonance. Furthermore, we predicted that under the resonance hypothesis, flicker and single pulse responses do not necessarily match, both in terms of presence and amplitudes. We already showed that a subset of contact locations (17.4-25.8%) had significant flicker responses without exhibiting single-pulse EP, indicating that single-pulse EP may not be necessary for steady-state EP (Figure 4C). We also found that the amplitudes of steady-state EP and single pulse EP are often different, with a tendency for stronger single-pulse EP (Figure 4D).

**Figure 5.**
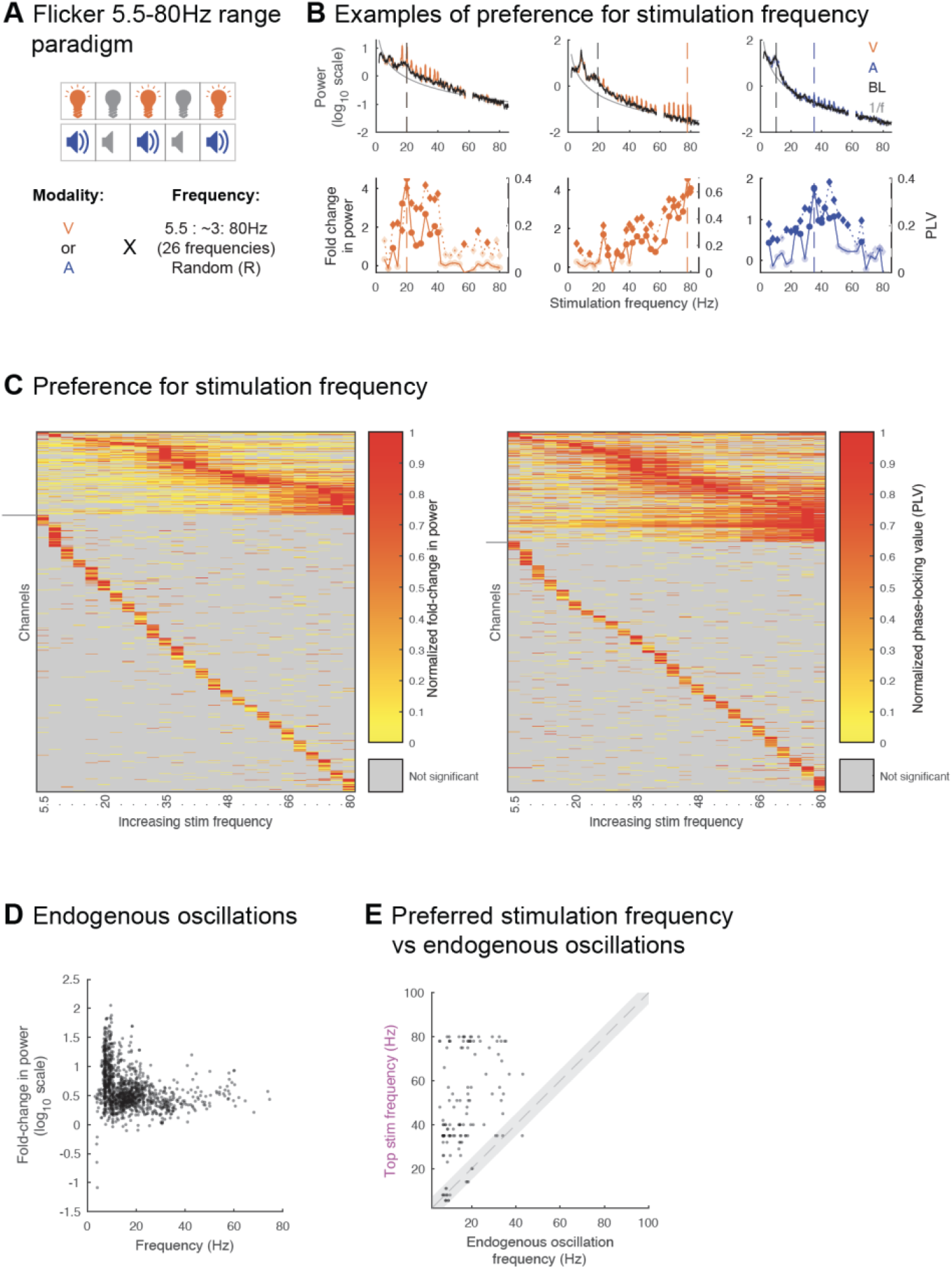
Flicker response is dependent on intrinsic circuit properties. (A) In the Flicker 5.5-80Hz range paradigm, subjects (5 subjects, 6 sessions) were exposed to either visual or auditory modalities, at 26 different frequencies spanning the 5.5-80Hz range, as well as random non-periodic flicker and baseline (no stimulation). (B) Example contacts (one example per column) showing endogenous oscillations and response to different stimulation frequencies. Top: power spectral density (PSD) during stimulation at each of the 26 flicker stimulation frequencies, only showing power values at the stimulation frequency +/- 1Hz for each condition overlaid on the average baseline PSD (black) and aperiodic fit (1/f, grey). Solid colored line indicates mean and shaded area indicates standard error of the mean. Bottom: corresponding fold-change in power (solid line) and phase-locking value (PLV, dotted line) for each stimulation condition. Vertical dashed colored line indicates stimulation frequency leading to maximal modulation. Vertical dashed grey line indicates frequency of detected endogenous oscillation that is closest to that top stimulation frequency. Solid discs and diamonds indicate significant fold-change and PLV, respectively. (C) Left: heatmap of normalized fold-change in power at the frequency of stimulation, for each channel (one channel per row) and frequency of stimulation (one frequency per column). Right: heatmap of normalized PLV for each channel and frequency of stimulation. For each channel, fold-change in power and PLV were normalized across the stimulation frequencies. Some channels are repeated for one subject, as that subject underwent both the visual and auditory versions of the Flicker 5.5-80Hz range paradigm. Channels are ordered from ones showing significant modulation to more than 6 stimulation frequencies (above horizontal grey lines to the left of the heatmaps), versus ones showing significant modulation to 6 or less stimulation frequencies (below horizontal grey lines), then by preferred or top stimulation frequency from lowest to highest. Channels with no significant modulation to any stimulation frequency are not represented. (D) Scatter plot of the fold-change in power relative to aperiodic fit, at the peak of each identified endogenous oscillation, versus the frequency of that endogenous oscillation, for all identified endogenous oscillations (see Methods) across all contacts from 5 subjects and across 6 sessions; each dot represents a contact in a session. (E) Scatter plot of the frequency of stimulation leading to maximal fold-change in power at the frequency of stimulation (top stimulation frequency) versus closest detected endogenous frequency for all contacts (across 5 subjects and 6 sessions) that showed at least one endogenous oscillation at baseline, and significant response to more than 6 of the flicker stimulation frequencies tested; each dot represents a contact in a session; grey shaded area represents +/- 5Hz from x=y.

Next, we tested whether flicker entrains endogenous oscillations. If flicker responses result from entrainment of endogenous oscillations, then flicker steady-state EP should be strongest at frequencies that are closest to those of endogenous oscillations^1^. To test this prediction, we compared the response to flicker stimulation at frequencies spanning the 5.5Hz-80Hz stimulation range, to baseline endogenous frequencies (see Methods). We found that across recorded contact locations, many showed strong endogenous oscillations in the alpha (∼10Hz) and beta (∼20Hz) ranges, with some also exhibiting higher frequency oscillations (Figure 5D). We hypothesized that stimulation frequencies eliciting maximal fold-change at the stimulation frequency, or top stimulation frequencies (examples illustrated with dashed colored line in Figure 5B), may be similar to strong endogenous oscillations. We focused on contacts with significant fold-change in power for more than 6 out of the 26 tested flicker frequencies because these tended to show clear preference for given frequency ranges, which may overlap with endogenous oscillations. Overall, among contact locations showing both clear baseline endogenous oscillations and significant steady-state EP for more than 6 out of the 26 tested flicker frequencies, we found only a minority (16.9%) of locations having the top stimulation frequency within 5Hz of an endogenous frequency (Figure 5E; paired, two-sided t-test of endogenous and optimal stimulation frequencies, p-value=0, ci=[-34.0664 -26.6039], tstat=- 16.0855, df=129, sd=21.5022). These results show that while flicker may entrain endogenous oscillations in specific cases, this does not explain most steady-state EP responses.

### Flicker decreases IEDs

Having established that flicker modulates widespread brain regions in humans, including deeper cortical structures and cognitive areas, we next evaluated whether flicker modulation alters the frequency of IEDs, epileptiform activity heavily involved in epilepsy and also implicated in several cognitive disorders including AD^50^. Of note, our recruitment criteria excluded subjects clinically suspected to be susceptible to photic-induced seizures, or who showed abnormal EEG activity in response to clinical photic stimulation (see Methods). Using an automated IED detection algorithm previously validated against clinical neurologists’ performance^67^ (see Methods), we identified and quantified the proportion of IEDs occurring during 10s trials of stimulation (Figure 6A) and their subsequent unstimulated baseline trials of the same duration. We found that for all conditions tested, on average across trials, a smaller proportion of IEDs occurred during stimulation (Figure 6B, left). Because we observed similar results across all stimulation conditions, we combined all conditions and computed a single proportion of IEDs during stimulation per session. Across conditions, flicker reduced IEDs in 10 out of 13 sessions across 12 subjects, by an average of 3.3% (range -12.9% to 6.0%, one-sample, one-sided t-test of a decrease from 50%, p-value=0.0234, ci=[-Inf 49.6792], tstat=-2.2152, df=12, sd=2.6720, Figure 6B right, and 6C). While there was an increase in IED rate in three sessions, and a small reduction in IED rate (<5%) in five sessions, five sessions show strong IED reduction (> 5%), suggesting that flicker may have larger effects in some individuals than others (Figure 6C). These results are unlikely to stem from the false positive rate of the IED detection algorithm, as we do not expect it to be biased towards any condition; if such bias were present, we would expect it to be towards stimulation conditions (i.e. increased false positives during stimulation due to sensory-evoked activity), which is opposite to our results. Overall, these results show that short 10s exposure to flicker modestly decreases IED rate, and that flicker stimulation does not constitute a risk of exacerbating IEDs in our patient population.

**Figure 6.**
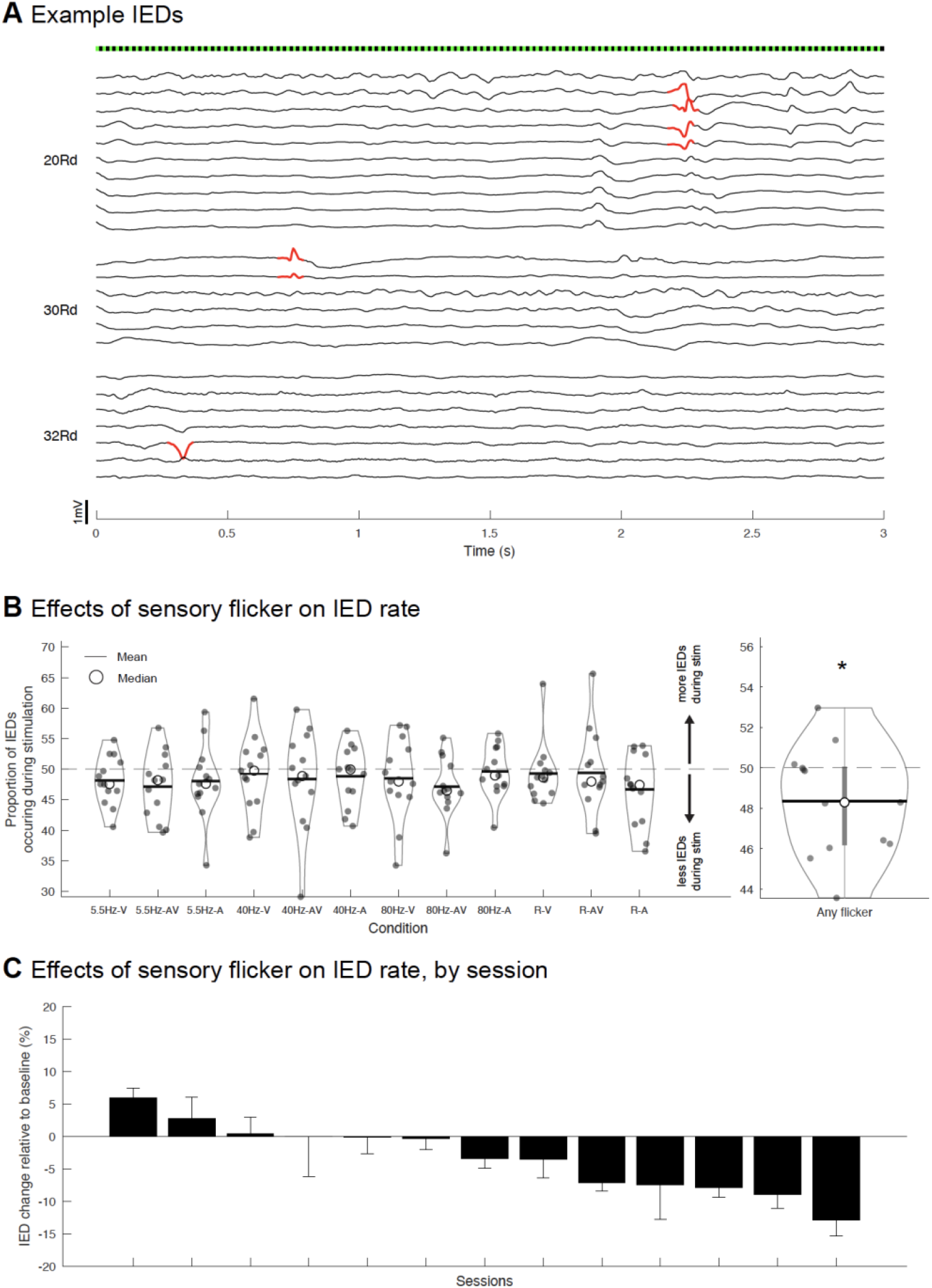
Decrease of interictal epileptiform discharge rate in response to flicker. (A) Example interictal epileptiform discharges (IEDs) detected (in red) over the first 3 seconds of a 40Hz-audiovisual (AV) trial, across 3 depth electrodes (labelled to the left as 20Rd, 30Rd and 32Rd) that had channels which detected those IEDs. Each trace represents preprocessed local field potential (LFP) signal from a contact of the depth electrode labelled to the left. (B) The proportion of IEDs detected during flicker stimulation versus the entire period (e.g., during the 10s of flicker stimulation versus 10s of flicker + 10s of subsequent baseline) for each condition and session (left). Each dot indicates one session (average over trials in the session, total of 13 sessions across 12 subjects). Because each condition showed the same trend (left), all conditions were combined to compute a single average per session (right). There was a significant (p-value<0.05, indicated by asterisk) decrease in the proportion of IEDs when combining all stimulation conditions. Open circles indicate median, horizontal lines inside violin plots indicate means, thick vertical line inside the violin plot on the right indicates range between the 25^th^ and 75^th^ percentiles. (C) Bar graph showing percent change in IEDs relative to baseline, by individual session, ordered from highest increase to highest decrease. Error bars represent standard error.

## DISCUSSION

Multisensory flicker stimulation was previously shown to modulate neural activity in rodent higher cognitive regions, and to provide therapeutic benefits with respect to Alzheimer’s disease-related neuropathology and cognitive outcomes^51,68^. Whether analogous flicker exposure can modulate human brain function to therapeutic effect, and by what mechanisms, was unclear. Accordingly, we studied the extent to which flicker affects brain activity in humans using invasive intracranial electrodes, the gold standard for localizing and characterizing neural activity. We found that flicker induces a steady-state EP across widespread brain networks, including canonical cognitive cortices in the MTL and PFC. Moreover, we observed that flicker-induced neural responses are consistent with the resonance of brain circuits but not linear superposition of single sensory pulse responses, offering a mechanistic explanation for previous observations and a more rational framework for optimizing stimulation parameters. Finally, we found that multisensory flicker acutely reduces the rate of IEDs, a biomarker of pathological activity implicated in epilepsy and other conditions including AD and autism.

By studying the neurophysiological effects of sensory flicker and single pulses with high spatiotemporal precision in extensively sampled cortices, we found that both steady-state EPs and single pulse EPs occur in many networks, within locally constrained neural populations. While steady-state EPs may be observed with non-invasive scalp EEG^25–35^, neural signals are highly filtered and attenuated with this recording method, with limited localization value. By contrast, intracranial electrode recordings provide improved localization. Indeed, intracranial electrode recordings have been used to show steady-state EP in the visual system, including the lateral geniculate nucleus, the primary and secondary visual areas^36^, and the broader occipital lobe^37^. We likewise used intracranial EEG, specifically SEEG, but showed that steady-state EPs occur in locally constrained neural populations across various circuits beyond canonical sensory regions. Three other studies^38–40^ have observed sensory-induced intracranial steady-state EP in insular, frontal, and temporal lobe structures. However, these studies were limited by either low sample size, restricted testing conditions, or poor referencing of the recordings which allowed for noise contamination or poor localization of the signal. Here, we systematically tested a wide spectrum of flicker conditions (12 to 27 stimulation conditions, in the different paradigms) including several frequencies and modalities, in a large set of subjects (16) and across widespread deep and superficial cortical regions. Moreover, we used Laplacian offline re-referencing of the recordings, a highly localizing method that minimizes volume conduction and noise^69^.

We found that flicker induces steady-state EPs throughout the sampled cortices, including regions associated with attention, limbic, and default-mode networks in addition to expected visual regions, as defined by resting state functional MRI studies, as well as regions involved in auditory processing. More specifically, we observed flicker-related responses in the MTL and PFC, showing the capacity of this intervention to non-invasively modulate brain structures involved in higher cognitive processes and in degenerative and other neuropsychiatric disorders. Although we observed significant steady-state EP in multiple cognitive regions, their amplitudes were typically lower than in sensory regions. This may be due to attenuation of the sensory signal as it is processed on the way to and through higher cognitive regions. Another potential factor is the stimulation parameter set used, including the intensity of stimulation (i.e., brightness and volume) or the optimal stimulation modality and frequency. For practical reasons, stimulation intensity was varied to each subject’s level of comfort (Table S4), and it is possible that higher intensities could produce even larger steady-state EPs in higher cognitive regions. The optimal stimulation intensities to modulate the MTL and PFC remain to be explored. We also note that, even with intracranial recordings, it can be difficult to disentangle whether a signal originates from truly higher cognitive processing, or early processing, such as the third visual pathway in the superior temporal sulcus^56^, or optic radiations lateral to the hippocampus^57^. We observed that some neural units in hippocampus and cingulate respond to flicker with higher average firing rates at a particular phase of the stimulus. Similar to our prior study in mice^51^, we did not expect all neurons to respond to a given sensory stimulus, since these are multisensory cognitive regions. Finally, although we showed clear modulation of the MTL and PFC, further work is needed to determine whether such modulation might have a meaningful clinical or functional impact on human memory. There is some evidence that sensory flicker can impact memory consolidation^14^, and current studies are pursuing potential therapeutic effects in AD. Our group’s prior study showed preliminary evidence that 8 weeks of sensory flicker strengthens functional connectivity of nodes in the default mode network in patients with AD^70^. Indeed, here we found flicker entrains locations that would be associated with multiple cognitive resting state networks, like the attention and default mode networks, that have weakened functional connectivity in AD. Flicker stimulation may affect functional connectivity by driving neurons within connected networks to be more likely to fire together^71^.

To understand the pathophysiological significance of sensory flicker, we assessed its effects on IEDs, a type of epileptiform activity. IEDs and other epileptiform activity are typical of epilepsy, but also may be observed in association with other neuropsychiatric disorders such as AD, multiple sclerosis, autism, and ADHD.^50^ In our cohort of subjects undergoing intracranial investigations of focal epilepsy, we observed that short (10s) duration sensory flicker exerts a modest anti-epileptic effect, acutely reducing IEDs. While rhythmic photic stimulation can elicit seizures in susceptible individuals^54^, patients with focal epilepsy are not generally susceptible to photic-induced seizures^72^. Moreover, our study recruitment criteria excluded patients clinically suspected to be susceptible to photic-induced seizures. Thus, sensory flicker was safe and possibly transiently therapeutic in our study population. Additional studies are required to optimize parameters, explore interactions with underlying brain states and disease conditions, and examine effects of more prolonged or repeated exposure to sensory flicker. These effects show that sensory flicker affects neural activity beyond generating steady-state EP and reduces a specific type of pathological neural activity.

Our study explored the mechanisms of steady-state EP using invasive intracranial recordings in humans. A prior study utilizing noninvasive scalp EEG^41^ suggested that the visual steady-state EP results from the linear superposition of single-pulse EPs. This proposed mechanism has been used to explain the observation that 40Hz auditory stimulation leads to a peak in response amplitude, with the 40Hz component of single-pulse EPs maximally summating at that frequency^32^. By contrast, our experimental paradigm using within-subject direct comparison between single-pulse EPs and steady-state EP contradicted linear superposition as a unifying mechanism. Importantly, we recorded local steady-state EPs intracranially in contrast to scalp EEG which measures summed and skull-filtered responses emanating primarily from early sensory structures that have the strongest responses. Another debated mechanism hypothesizes that flicker entrains endogenous oscillations. One study^45^ used magnetoencephalography (MEG) to show that visual cortex gamma oscillations (induced by viewing a grating stimulus) are not entrained by visual flicker. The authors expected an increased steady-state EP amplitude when stimulating at a frequency close to the subject’s intrinsic gamma frequency of the visual system. Instead, they observed a steady decrease in response amplitude as a function of increasing stimulation frequency, regardless of the proximity to intrinsic gamma. In contrast, we found many channels across subjects showed a stronger steady-state EP for specific stimulation frequency ranges, with only a minority showing a decrease in amplitude as a function of increasing stimulation frequencies. These divergent findings may be due to our directly sampling beyond visual areas, or a higher signal-to-noise and spatial resolution of our recordings compared to MEG. Moreover, we found that for a minority of contacts, optimal stimulation frequencies resembled baseline endogenous oscillation frequencies.

Instead of linear superposition of single sensory pulse responses, we find that flicker-induced neural responses are consistent with the resonance of brain circuits. Our study showed that putative circuits resonate to specific flicker stimulation frequencies, with many recording locations exhibiting a peak response to stimulation in specific narrow frequency ranges. These findings are consistent with a prior study^12^ in which optogenetic stimulation of fast-spiking interneurons in the mouse sensory barrel cortex at various frequencies led to the highest increase in LFP power in the gamma range, illustrating an optimal resonant property of that cortical circuit. Such resonance may depend on the cellular composition of the circuit^62^, the nature of recurrent synapses, biophysical properties of local and input neurons, and modulatory input from other circuits. Together, these results illustrate the importance of selecting an optimal frequency to maximize steady-state EP in target regions, which will require further characterization and understanding optimal resonance of neural circuits.

Despite its novel findings regarding the neurophysiology of flicker and its potential therapeutic benefits in humans, this study has limitations. First, our subjects’ brains were variable, often harboring brain lesions from previous accidents or surgeries, or various pathologies (Table S2). Patients varied by age, sex, cognitive deficits, and baseline seizure medication regimens, although this is partly mitigated by widespread brain sampling and a relatively large group of subjects. Second, the brain regions sampled across our patients undergoing intracranial monitoring of focal epilepsy were by nature enriched for regions suspected and/or proven to harbor pathological seizure networks, raising the possibility that observations might be specific to epileptic rather than healthy brains. While this potential confound cannot be eliminated, widespread sampling included brain regions that were ultimately found to be outside seizure onset zones or abnormal regions: across our 16 subjects, the majority of contacts were outside abnormal tissue and/or seizure onset zones. Third, the clinical environment in which experiments took place involved factors related to the clinical care of the subjects including varying environmental stressors, postoperative discomfort, sleep deprivation, changing medication dosages, and other factors which could affect brain states.

This study bridges findings from rodents to humans and shows that multisensory flicker non-invasively modulates brain circuits, including those involved in higher cognitive processes and impacted in disease, potentially to therapeutic effect. This investigation is unique in using extensive direct intracranial neural recordings in humans to characterize single-pulse and steady-state EP with high spatiotemporal resolution. Furthermore, we elucidated the mechanisms of steady-state EP in multiple circuits, shedding light on strategies to maximize modulation. Finally, our findings demonstrate proof of concept that flicker can reduce IEDs, a pathological brain activity associated with epilepsy, AD, autism, and other disorders.

## METHODS

### Participants

We recruited treatment-resistant epilepsy patients undergoing pre-surgical intracranial seizure monitoring. In between clinical services and at the patient’s discretion, one or more of three experimental paradigms (Figure 1B, Figure 4B, and Figure 5A) were carried out in the patient’s room. All study-related procedures were approved by the Emory Institutional Review Board. The study was registered at clinicaltrials.gov under NCT04188834. Recruitment criteria included: age over 18, fluent in English, able to understand and give verbal and written consent to the study procedures and associated risks, not suspected to be susceptible to photic-induced seizures or psychogenic non-epileptic seizures (PNES) triggered by sensory stimulation, did not show abnormal EEG activity if tested with clinical photic stimulation. We obtained informed consent from all recruited subjects, and 16 of the recruited subjects completed (15 subjects) or neared completion (1 subject) one or more paradigms overall (Table S4).

### Electrophysiology

As part of their clinical planning, patients were implanted by a neurosurgeon (JTW or REG) with SEEG depth electrodes, most often from DIXI Medical (DIXI Medical, France), with 0.8mm diameter, 2mm length platinum/iridium contacts, typically separated by 3.5mm intervals center-to-center. In some subjects, depth electrodes included (usually 1-2 per candidate, consenting subject) FDA-approved Ad-Tech electrodes (Ad-Tech Instrument Corp, Racine, WI; 1.28mm diameter, 1.57mm length platinum contacts, separated by 5mm intervals center-to-center) containing nine 38-micron microwires protruding from their tip that allowed recording from single neurons. The number and implant location of the stereotactic depth electrodes were exclusively determined by the clinical team and based on clinical needs. Local field potentials measured with macro-contacts were recorded on the clinical monitoring system (XLTEK EMU 128FS; Natus Medical) and associated Natus Neuroworks software, typically at a rate of 2,048Hz (range 1,024-16,384Hz). The clinical system’s reference and ground were typically sub-galeal contacts from an electrode array placed sub-dermally at the vertex. Microwires were recorded using the Blackrock NeuroPort system (Blackrock Microsystems, UTSW) and associated Central Software Suite at a rate of 30,000Hz using a dedicated microwire as physical reference.

### Stimulus presentation

Customized software ran in MATLAB 2019b controlled stimulus presentation, including the creation of analog sensory signals and pulses synching the EEG recordings, and control of their timing (a version of the source code is available at https://github.com/singerlabgt/Behavioral_FlickerMasterTask). These signals were produced using a digital acquisition board (USB-6212 multifunction I/O device, National Instruments), which sent analog signals to a customized circuit. Opaque glasses containing LEDs (Mind Alive Inc.) administered visual stimuli, and earbuds (SONY MDR-EX15LP) presented auditory stimuli. These glasses maximized the extent of the visual field stimulated, while earbuds maximized signal of auditory stimuli relative to surrounding noise. Visual stimuli consisted of a 50% duty cycle square wave signal, while auditory stimuli consisted of a 7kHz tone amplitude-modulated by a pseudo-square wave envelope, with about 1.6ms ramp up and down each to minimize noise due to amplifiers rapidly turning on and off with each cycle. We opted to use a 50% duty cycle square wave signal for sensory stimuli, as it was previously shown that such a visual square wave signal had the highest likelihood of inducing a steady-state EP in the occipital cortex, compared to other types of waves such as triangular or sinusoidal^73^. Stimulation trials were synchronized with neurophysiology acquisition systems via a TTL pulse.

At the start of any experiment, we first adjusted the brightness and volume to each subject’s comfort, then administered individual trials from each sensory stimulation condition to check for any evidence of associated seizure symptoms and/or activity. To control for the possibility that stimulus responses were due to an artifact from the sensory stimulation apparatus, participants underwent a relative occluded condition in which stimuli were delivered but occluded with a sleep mask and/or towels on their eyes underneath the glasses, and earplugs while earphones were near but not placed into the ears. At the end of each session, we measured the brightness and volume of the device at 40Hz audiovisual stimulation, using a luxmeter (TRACEABLE Dual-Range Meter) and decibel meter (BAFX Products BAFX3608).

Subjects were exposed to one or more of the following three stimulation paradigms (Figure 1B, Figure 4B, and Figure 5A):

#### 1) Flicker 5.5-40-80Hz paradigm

Subjects were exposed to 10-second trials including modalities of visual only, audio only, and audiovisual combined, at frequencies with 50% duty cycle of 5.5Hz, 40Hz, 80Hz, and random pattern. Trials were pseudo-randomized (with no given modality or frequency repeated more than three times in a row) to control for order effects and minimize habituation to a given stimulus. Each stimulation trial was followed by 10 seconds of no stimulation, i.e. a baseline trial. In the audiovisual conditions, light and sound onset and offset were synchronized. Random pattern stimulation consisted of 12.5ms pulses with inter-pulse intervals randomized for durations between 0-25ms (i.e. average light exposure duration per period and average frequency was around those of the 40Hz conditions). In total, over the about 1h-long experiment, subjects were exposed to 360 10-second trials, with 15 trials per stimulation condition, and 180 no-stimulation (i.e. baseline) trials. During the experiment, subjects were asked to keep their eyes open, in order to maximize the visual steady-state EP, and offered breaks every 10 minutes. The relative occluded condition typically consisted of 6 10-second 40Hz-visual trials and 6 10- second 40Hz-audio trials.

#### 2) Single-pulse paradigm

To compare flicker responses to those generated by single pulses of light or sound, subjects were exposed to 12.5ms-long single pulses of visual only, audio only, or audiovisual modality, with inter-pulse intervals randomized between 987.5-1487.5ms. The single pulses were identical in duration to the 40Hz flicker condition. Each condition was repeated for a total of 200 trials. Trials from each modality were presented in a pseudorandomized manner (no given modality repeated more than three times in a row). The relative occluded condition consisted of 200 audiovisual trials.

#### 3) Flicker 5.5-80Hz range paradigm

To map flicker responses across a wide range of frequencies, subjects were exposed to 10- second trials of either visual or auditory modality, at frequencies spanning the 5.5Hz-80Hz range: 5.5Hz, 8Hz, 11Hz, 14Hz, 17Hz, 20Hz, 23Hz, 26Hz, 29Hz, 32Hz, 35Hz, 38Hz, 40Hz, 42Hz, 45Hz, 48Hz, 51Hz, 54Hz, 57Hz, 63Hz, 66Hz, 69Hz, 72Hz, 75Hz, 78Hz, and 80Hz. In cases where only one session of a given modality was administered, the modality was typically picked based on individual subject’s electrode coverage, i.e. visual modality was selected in subjects with electrodes in areas predicted to respond to visual stimulation more than auditory stimulation. Each condition included 10 trials. We also included 10 trials of random pattern stimulation and 10 trials of no stimulation (baseline). Trials were pseudo-randomized (no given condition repeated more than three times in a row, with attempt to spread all conditions across the experiment duration) and separated by an intertrial interval randomized between 2-2.5s. Random pattern stimulation and flicker duty cycle were the same as in the Flicker 5.5-40-80Hz paradigm. Subjects were asked to keep their eyes open and offered breaks about every 10 minutes. The relative occluded condition consisted of 6 10-second 40Hz trials of the selected modality.

### Electrode localization

As part of stereotactic planning and confirmation, subjects typically received structural T1 and T2 MRIs before the electrode implant surgery, and a CT scan and a structural T1 MRI following surgery. We identified and labelled electrodes on the post-operative CT using the voxTool software (https://github.com/pennmem/voxTool), then co-registered all imaging to pre- operative T1 MRI using rigid transformation with the Advanced Normalization Tools package (ANTs; stnava.github.io/ANTs/;^74^). We calculated electrode coordinates in different imaging spaces using co-registration output and custom MATLAB scripts that incorporated a function from Lead-DBS (lead-dbs.org;^75^). Pre-operative T1 MRI was parcellated and segmented using the FreeSurfer toolbox (https://surfer.nmr.mgh.harvard.edu/;^76^). Where appropriate, here and in other preprocessing steps or analyses, we used GNU Parallel to process data in parallel^77^ (https://www.gnu.org/software/parallel/). Electrodes were anatomically labelled using FreeSurfer outputs and custom scripts. Anatomical label assignment was performed to identify the label from which the electrode was likely picking up the strongest signal, using a 5mm radius gaussian search sphere centered at the center of mass of the electrode. Specifically, we 1) converted the center of each atlas voxel to RAS world coordinates, 2) searched for all anatomical labels within the search sphere around the electrode’s center of mass with a radius r of 5mm, 3) removed atlas voxels labeled as cerebral white matter or white matter hypointensities, 4) for these remaining voxels, assigned a signal strength amplitude, calculated as 1/r (for r=0, assigned value of 1), 5) summed labels and associated signal strength, 6) picked the label with the greatest overall signal strength. We extracted normalized electrode locations into MNI space, through rigid, affine then symmetric image normalization (SyN) coregistration of pre-operative T1 MRI to T1 MNI MRI (ICBM152 2009b Nonlinear Asymmetric;^78,79^). Where brain imaging was used to show electrode location (Figure 1 and Figure 2), where appropriate we rotated the imaging volumes to the plane of the depth electrode of interest, in order to clearly see all contacts from the electrode.

### Identification of pathological features of recorded locations

To assess the proportion of results originating from recording locations involved in abnormal tissue or seizure network (see Discussion), we used the subjects’ clinical reports, including imaging and neurophysiological reports. We identified contacts located in or near abnormal tissue (such as previous resection, encephalopathy, or periventricular nodular heterotopia), the seizure onset zone(s), and contacts implicated in the detection of interictal epileptiform discharges. When estimating the percentage of recording locations in or near abnormal tissue or seizure onset zone(s), we included any recording site which Laplacian montage included any channel tagged with such features.

### Data analysis

Most analyses were run in MATLAB 2019b, using custom scripts in combination with Fieldtrip^80^ (https://www.fieldtriptoolbox.org/) and Chronux (http://chronux.org/) toolboxes.

#### Exclusion of LFP channels from analysis

We identified contacts outside brain parenchyma, in CSF, in ablation or resection lesion or cavity, and contacts that were defective or showing artifacts based on the clinical team anatomical labelling of electrodes and neurophysiology reports. In cases where anatomical labelling from the clinical team was not available, contacts outside brain parenchyma or in CSF were manually identified using preop T1 MRI coregistered to postop CT. Moreover, for each experimental session noisy channels were identified by visual inspection of signals from a randomly selected set of 10-second segments, in both the time and frequency domains. All above channels were excluded from further preprocessing and analysis.

#### Referencing of LFP recordings

In most analyses, we aimed to localize any sensory response to the best approximation of their neurophysiological source. The monopolar referencing provided by the clinical recording system has several disadvantages, including signal and noise contamination from the subdermal physical reference placed at the vertex, and being prone to volume conduction^69^. We used Laplacian re-referencing of the LFP recordings for all analyses, as it was shown to be optimal at localizing the source of recorded signals^69,81^. The Laplacian montage is a highly localizing method, whereby volume-conducted signal across multiple contacts is minimized, while local signal is preserved. Moreover, noise resulting from movement artifact or ground noise is also greatly reduced, providing a cleaner signal. This referencing method takes the signal from each contact and subtracts the average from the two most adjacent contacts. For contacts at the extremities of depth electrodes, bipolar referencing was used (i.e. the signal from the adjacent contact was subtracted from the signal from the end contact). In cases where a channel was adjacent to an eliminated channel (due to it being noisy for instance, see above), we approximated Laplacian referencing by subtracting the mean from the two most adjacent contacts that were not eliminated. We treated bipolar referencing of end contacts with adjacent eliminated contacts similarly.

#### Processing and quantification of LFP response to flicker

For paradigms involving flicker stimulation (i.e. Flicker 5.5-40-80Hz and Flicker 5.5-80Hz range), LFP recordings were segmented into 12-second segments corresponding to each 10- second trial +/- one second to manage filter artifacts in a subsequent preprocessing step. For experimental sessions where there was an unequal number of trials per condition (for instance in case where the session was stopped before completion, or in the case of baseline trials for the Flicker 5.5-40-80Hz experiment), a random subset of trials was selected for analysis for those conditions with a higher number of trials, so that all conditions had the same number of trials. Data were re-referenced as detailed above and bandpass filtered between 2-300Hz, with a baseline correction over the duration of the 12-second segments. Power spectral density (PSD) was calculated for each 10s trial, over 2-100Hz, using the Chronux toolbox (http://chronux.org/), with a time-bandwidth product of 3, and number of tapers of 5.

To compare the amplitude of the steady-state EP across contacts, conditions, and subjects, for each contact and flicker condition we quantified the normalized fold-change in power at the frequency of stimulation using the following equation (similar to prior work^45^):

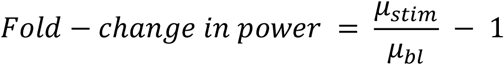

where μ_stim_ is the power at the frequency of stimulation averaged across trials from a given condition, and μ_bl_ is the power at the frequency of stimulation averaged across an equal number of trials from the baseline condition. For each contact and condition, we also calculated a corresponding significance value of the steady-state EP, using the following method: we computed μ_stim_ - μ_bl_, then performed a random permutation test of trial values with 10,000 iterations; cases where less than 5% of the resulting putative differences of the means were greater than the measured difference of the means, were considered significant. For illustration of the flicker response in the time domain, each 10s trial of a given condition was re-segmented into 2 cycles of the stimulus, with one overlapping cycle between consecutive segments. The resulting segments were averaged, and the standard error of the mean was calculated.

#### Processing and quantification of phase-locking value

For each contact and flicker condition, we calculated the inter-trial phase-locking value (PLV) to the stimulus, as in previous studies^45,82^. Specifically, the sensory stimulation signal was approximated with a sinusoid, then for each trial we calculated the cross-spectrogram (MATLAB’s function xpectrogram) of it and the preprocessed LFP signal, with window of size half the sampling frequency, and no samples of overlap between segments. The angle difference between the two signals was then calculated for each timepoint, then averaged across trials and time, and the absolute value was used as the PLV. The significance of the angle difference across trials was assessed using the p-value of Rayleigh’s test for non-uniformity of circular data, calculated using a circular statistics toolbox^83^. P-values below 0.05 were considered significant.

#### Detection of baseline endogenous oscillations

We measured the mean frequency and amplitude of endogenous oscillations at each recording location at baseline (no stimulation). To account for the aperiodic component of the PSD, which can influence the measures of endogenous oscillations, we used the FOOOF toolbox^84^ (https://github.com/fooof-tools/fooof). We used the following detection parameters on the mean baseline PSD for each recording location: max number of peaks of 5 per location, peak width limits between 2-10Hz, minimum peak height of 0.6, and frequency range of 2-100Hz. We then quantified the amplitude of each detected endogenous oscillation by fold-change in power of the modeled PSD at the center frequency of a given oscillation relative to the aperiodic fit of the PSD.

#### Processing of LFP single-pulse evoked response

Recordings from the duration of the experiment +/-60s were re-referenced, high-pass filtered at 0.1Hz (Butterworth IIR filter, order 4), segmented into 1s trials +/-0.25s, and baseline corrected for 0.25s before the start of trial. We then calculated the time-locked average and standard error of the LFP segments across trials. We evaluated whether a contact showed single pulse response by subtracting the root mean square of the averaged LFP from 0 (start of 12.5ms pulse) to 1s for the relative occluded condition from that of the stimulation condition, and performing a random permutation test of the trials with 500 iterations; cases where less than 5% of the resulting putative differences of the root mean square of the means were greater than the measured difference, were considered significant. We quantified the amplitude of the response by taking the absolute maximum peak of the response from the onset of the stimulus to 1s after the onset of the stimulus. To compare the amplitude of the flicker response versus single-pulse response (Figure 4D), we normalized the response to a given stimulus type (flicker or single pulse) by modality for each subject from 0.001 (minimum value) to 1 (maximum value), then took the log_10_ of those values.

#### Anatomical characterization of the sensory response

To describe the anatomical location of the response to sensory stimulation across brain regions and subjects, we adopted three different strategies. First, we represented the location of recording LFP contacts in 3-dimensionsional normalized space with associated size and color codes representing whether modulation was significant and amplitude of the response, respectively. Second, we assessed anatomical regions based on FreeSurfer-outputted anatomical labels. Finally, we plotted a heatmap of the sensory response as a function of assigned anatomical label of the electrode, and by condition and subject. For most analyses, the FreeSurfer provided Desikan-Killiany parcellation atlas and Fischl et al. 2002^85^ segmentation atlas labels were used. For Figure 1D and 1F, the visual group included FreeSurfer labels pericalcarine, cuneus, lingual and lateral occipital, while the auditory group included transverse temporal and superior temporal. For Figure 2C and 2D, the MTL group included FreeSurfer labels temporal pole, amygdala, hippocampus, entorhinal and parahippocampal, while the PFC group included medial orbitofrontal, rostral anterior cingulate, caudal anterior cingulate, frontal pole, superior frontal, rostral middle frontal, caudal middle frontal, lateral orbitofrontal, pars orbitalis, pars triangularis and pars opercularis. For analyses involving functional networks, we used surface-to-surface coregistration of Yeo 2011’s labels^59^ provided by FreeSurfer in their fsaverage space, to individual subjects. These labels correspond to a set of 7 networks clustered via resting state functional connectivity across 1,000 healthy subjects. Here, and elsewhere, final figure panels were outputted in part using code from the export_fig toolbox (https://github.com/altmany/export_fig). Moreover, violin plots were produced using code from the Violinplot-Matlab toolbox^86^ (https://github.com/bastibe/Violinplot-Matlab).

#### Processing of neuronal unit response to sensory stimulation

For each microwire recording, spikes were extracted and clustered using the Combinato Python-based software^87^ (https://github.com/jniediek/combinato), with threshold for extraction 6 times the standard deviation of noise. We then manually classified outputted groups of sub- clusters as artifact, putative multi-unit, or putative single neuron using criteria similar to previously defined^88^. Mainly we used the following strategy:

**Table 1:** Criteria for classifying a group as artifact, multi-unit or single-unit.

| Description | Artifact | Multi-unit | Single-unit |
| --- | --- | --- | --- |
| Average waveform shapes (visual inspection) for all unit sub-clusters (can be helped by plotting of all waveforms) | Looks like artifact | Does not look like artifact | Does not look like artifact |
| Firing rate (per second) | $\leq 0.05$ | $> 0.05$ | $> 0.05$ |
| Fraction of inter-event intervals $< 3\text{ms}$ | $\geq 0.1$ | $\geq 0.05$ | $< 0.05$ |
| Narrowing of main peak of waveforms' density plot (visual inspection) | Narrowing | Narrowing | No narrowing |
| Local peaks (positive or negative) following main peak of average waveforms (visual inspection) for all sub-clusters | $\geq 4$ | 3 | $< 3$ |
| Distribution of the event amplitudes (visual inspection) | - | Multimodal | Unimodal |

A group was classified as a single unit if it satisfied all criteria, as a multi-unit if it did not meet any of the artifact criteria, and as an artifact otherwise. Moreover, a group was considered an artifact if events tended to occur within confined periods of the experiment.

For analysis of the effects of flicker on spiking activity, ten second trial segments were re-segmented into two stimulus cycles, with one cycle overlapping between pairs of cycles. In the case of random flicker condition, cycles were composed of adjacent 25ms segments (again, with one overlapping segment between pairs of cycles). For each unit, we then calculated the peristimulus-time histogram aligned to the onset of each pair of stimulus pulses. Units with no spikes for more than 20% of the peristimulus time histogram bins in all conditions were deemed to have too few spikes to assess whether they were modulated by sensory flicker, and thus were eliminated from further analyses. To determine the strength of spiking modulation by stimulus phase of each unit, we calculated the vector strengths and Rayleigh statistics of units for each stimulus condition using a circular statistics toolbox^83^.

#### Modeling of the linear superposition of single pulse evoked potentials

We modeled the linear superposition of single pulse evoked responses by generating simulated flicker responses from summed single pulse responses. We generated 15 10s 40Hz simulated flicker trials (visual, audiovisual, and auditory) by linearly summing randomly selected (among 200 trials) single-pulse EPs every 25ms. This is analogous to the 40Hz flicker stimulation, with 12.5ms pulses repeated every 25ms. Using randomly selected trials (as opposed to the averaged single pulse response) accounted for variability of the response to single-pulses. The amplitude of the simulated flicker response was then calculated in the same way as for the recorded flicker response.

#### IED detection and analysis

Manual detection of IEDs is a time-consuming process requiring expert clinical input. We thus opted to use a previously validated automated IED detection algorithm^53,67,89^. Channels eliminated for the flicker modulation analysis (because of being outside brain parenchyma, noisy, or other reasons mentioned above) were also eliminated before performing preprocessing and IED detection. Spikes occurring within a time window of 100ms were considered part of the same IED (i.e. a polyspike), and IEDs detected across more than 11 channels were deemed to be noise and eliminated, as was done in a previous study^53^. The frequency of detected IEDs was generally highly variable on the timescale of minutes. To control for this variability, we compared the frequency of IEDs during stimulation trials with their directly subsequent baseline trials (both stimulation and baseline trials occurring within 20s). Specifically, for each trial the proportion of spikes occurring during the 10s of stimulation, out of the whole trial duration (stimulation and following baseline period), was calculated, then averaged within each condition. When averaging, we omitted 20s trials where no IEDs were detected. Given that we observed a similar trend of a decrease in IEDs during stimulation compared to baseline across all stimulus conditions, we averaged the stimulation IED proportion across conditions, obtaining one IED proportion value for each session. Two of the sessions’ results came from one subject. Since they differed, with one session showing reduction in IEDs during stimulation, and one session showing increase in IEDs during stimulation, they were treated as independent.

## Data Availability

De-identified, minimally processed data and processed data, will be made available upon request.

## Data Availability

De-identified, minimally processed data and processed data, will be made available upon request. Corresponding authors will respond to requests within two weeks of receipt.

## Code Availability

Code used for preprocessing and analysis of the data will be made available on GitHub upon publication.

## ACKNOWLEDGMENTS

First, we would like to thank both patients and clinical staff of the Emory University Hospital Epilepsy Monitoring Unit for their participation and assistance with the study. A.C.S. acknowledges the Packard Foundation and NIH NINDS R01 NS109226; J.T.W. acknowledges NIH NIMH R01 MH120194-01A1. LTB was in part supported by DARPA-BAA-14-08, U01NS113198, NIMH R01 MH120194-01A1 and the Georgia Tech/Emory NIH/NIBIB Training Program in Computational Neural-engineering (T32EB025816). We thank members of the Singer laboratory and National Center for Adaptive Neurotechnologies (NCAN) at Washington University, as well as L.B. for feedback on the methods, results, and manuscript.

## AUTHOR CONTRIBUTIONS

LTB, JTW, and ACS designed the study; LTB performed research and analyzed the data; EC and JP assisted in localizing electrode contacts; MYW built the circuit for the sensory stimulation device; J.T.W and R.E.G. implanted electrodes in subjects; B.T.C consulted on IRB protocol and recruitment of candidate subject, especially in terms of their risk for photic-induced seizures; LTB, JTW, and ACS wrote the paper.

