## Supplementary Information for "Multisensory Flicker Modulates Widespread Brain Networks and Reduces Interictal Epileptiform Discharges in Humans"

**for**


### SUPPLEMENTARY TABLES:

| Subject | Sex | Age | Language dominance (as determined by fMRI) |
|---|---|---|---|
| **01** | F | 26-30 | L |
| **02** | M | 31-35 | L |
| **03** | F | 46-50 | L |
| **04** | F | 26-30 | L |
| **05** | F | 46-50 | L |
| **06** | F | 21-25 | L |
| **07** | M | 21-25 | L |
| **08** | M | 51-55 | B |
| **09** | M | 36-40 | L |
| **10** | M | 26-30 | L |
| **11** | F | 21-25 | R |
| **12** | F | 31-35 | L |
| **13** | F | 26-30 | L |
| **14** | F | 21-25 | L |
| **15** | M | 31-35 | L |
| **16** | F | 21-25 | L |

*Table S1. Subject demographics*

*M - male; F - female; R - right; L – left; B – bilateral.*

| Subject | Prescribed AEDs | AEDs on day of testing | Preoperative imaging findings | Determined seizure focus |
|---|---|---|---|---|
| **01** | Zonisamide, lamotrigine, levetiracetam | Lamotrigine, levetiracetam, zonisamide | History of left hippocampal sclerosis, expected post-operative findings of mesial temporal ablation. | Left temporal |
| **02** | Lamotrigine, levetiracetam, topiramate | Lamotrigine, levetiracetam, topiramate | History of prior left medial occipital-parietal resection, possible bilateral hippocampal sclerosis. | Right temporo-occipital region |
| **03** | Eslicarbazepine, lamotrigine | Lamotrigine | Likely left hypothalamic hamartoma. | Bilateral medial temporal |
| **04** | levetiracetam, lamotrigine, lorazepam, gabapentin | Flicker 5.5-40-80Hz session: lamotrigine, levetiracetam Single-pulse session: none | Question of medial left temporal cortical displasia. | Left medial temporal |
| **05** | Clobazam, levetiracetam, phenytoin. | Flicker 5.5-40-80Hz session: none Single-pulse session: levetiracetam, lorazepam | Right hemispheric atrophy, right mesial temporal sclerosis. | Right temporo-occipital region |
| **06** | Lamotrigine, lacosamide, perampanel. | Flicker 5.5-40-80Hz session: lamotrigine Single-pulse session: lamotrigine | Expected post-operative findings of left medial temporal ablation. | Left posterior parahippocampal area |
| **07** | Topiramate, phenytoin, gabapentin, clonazepam | Flicker 5.5-40-80Hz session: phenytoin Single-pulse session: none | Left frontal lobe polymicrogyria and associated closed-lip scattered schizencephaly. | Left fronto-parietal region |
| **08** | Lacosamide, lamotrigine | Lacosamide, lamotrigine | Expected post-operative findings of left temporal pole ablation. | Left orbitofrontal region |
| **09** | Lamotrigine | Flicker 5.5-40-80Hz session: lamotrigine Single-pulse session: none | Possible anterior right frontal focal cortical dysplasia. | Left mesial temporal |
| **10** | Levetiracetam, zonisamide, Lamotrigine. | Flicker 5.5-40-80Hz session: lamotrigine Single-pulse session: lamotrigine, levetiracetam | Possible right hippocampal atrophy. | Bilateral medial temporal |
| **11** | Lacosamide | None | Bilateral occipital periventricular nodular heterotopia. | Right parieto-occipital region |

| | | | | |
|---|---|---|---|---|
| **12** | Levetiracetam, lamotrigine | Flicker 5.5Hz-80Hz range session: none<br>Flicker 5.5-40-80Hz session 1: lamotrigine, levetiracetam<br>Flicker 5.5-40-80Hz session 2: none | No abnormal findings | Left medial temporal |
| **13** | Levetiracetam, zonisamide | None | No abnormal findings | Left basal/lateral temporal |
| **14** | Clobazam, lamotrigine, perampanel | Clobazam, lamotrigine | Small left frontal white matter cavernous malformation with associated developmental venous anomaly. | Left posterior frontal (perirolandic) |
| **15** | Carbamazepine, levetiracetam | None | History of radiosurgery of left temporal lobe and frontal operculum arteriovenous malformation. | Left planum temporale, Heschl's gyrus, pars opercularis. |
| **16** | Levetiracetam, lamotrigine | Flicker 5.5-80Hz range session 1: lamotrigine (100mg).<br>Flicker 5.5-80Hz range session 2: levetiracetam (250mg). | History of left temporal lobe low-grade (WHO grade 1) tumor, bilateral gray matter heterotopias, hypothalamic hamartoma, retrocerebellar cyst. | Poorly localized, multifocal; onset possibly left mesial temporal, bilateral, or multifocal |

*Table S2. Epilepsy information for each subject*
*AED – anti-epileptic medication.*

| Subject | IED rate (IED/min) | Seizure events | | |
|---|---|---|---|---|
| | | clinical | subclinical | total |
| **01** | 15.7 | 22 | 6 | 28 |
| **02** | 122.7 | 1 | + | 1+ |
| **03** | 79.9 | 13 | 4 | 17 |
| **04** | 32.6 | 5 | 0 | 5 |
| **05** | 61.0 | 11 | 20 | 31 |
| **06** | 30.0 | 5 | 1 | 6 |
| **07** | 9.6 | 5 | 0 | 5 |
| **08** | 54.5 | 4 | 0 | 4 |
| **09** | 36.6 | 7 | 0 | 7 |
| **10** | 44.5 | 4 | + | 4+ |
| **11** | n/a | 11 | 0 | 11 |
| **12** | 47.4 | 2 | 0 | 2 |
| **13** | 25.4 | 1 | 0 | 1 |
| **14** | n/a | 17 | 1 | 18 |
| **15** | n/a | 2 | 1 | 3 |
| **16** | n/a | 21 | 1 | 22 |

*Table S3. Intracranial monitoring activity per subject*

*IED – interictal epileptiform discharge; n/a – not available; + – multiple, not counted. IED rate was based on number of IEDs detected over the duration of the Flicker 5.5-40-80Hz experimental session.*

| Subject | Paradigm | | | | | |
|---|---|---|---|---|---|---|
| | Flicker 5.5-40-80Hz | | Single-pulse | | Flicker 5.5-80Hz range | |
| | Brightness (Lux) | Volume (dbA) | Brightness | Volume | Brightness | Volume |
| **01** | 163 | 76 | n/a | n/a | n/a | n/a |
| **02** | 199 | 82 | n/a | n/a | n/a | n/a |
| **03** | 14 | 83 | n/a | n/a | n/a | n/a |
| **04** | 49 | 78 | 715 | 93 | n/a | n/a |
| **05** | 136 | 72 | 1029 | 96 | n/a | n/a |
| **06** | 815 | 72 | 1003 | 74 | n/a | n/a |
| **07** | 122 | 80 | 162 | 96 | n/a | n/a |
| **08** | 1125 | 80 | n/a | n/a | n/a | n/a |
| **09** | 978 | 89 | 970 | 93 | n/a | n/a |
| **10** | 880 | 84 | 1063 | 100 | n/a | n/a |
| **11** | n/a | n/a | n/a | n/a | 158 | n/a |
| **12** | 212 | 79 | n/a | n/a | 233 | n/a |
| **13** | 189 | 77 | n/a | n/a | n/a | n/a |
| **14** | n/a | n/a | n/a | n/a | n/a | 85 |
| **15** | n/a | n/a | n/a | n/a | n/a | 95 |
| **16** | n/a | n/a | n/a | n/a | 82 | 79 |
| **Total** | 12 | | 6 | | 5 | |

*Table S4. Paradigm and sensory stimulation amplitudes per subject*
*A total of 16 subjects completed one of more of three paradigms (see Figure 1B, 4B, and Methods for details). For each subject and paradigm, participation as well as brightness and volume (averaged between left and right sides of the glasses or earbuds) measured at 40Hz are indicated.*

**SUPPLEMENTARY FIGURES:**

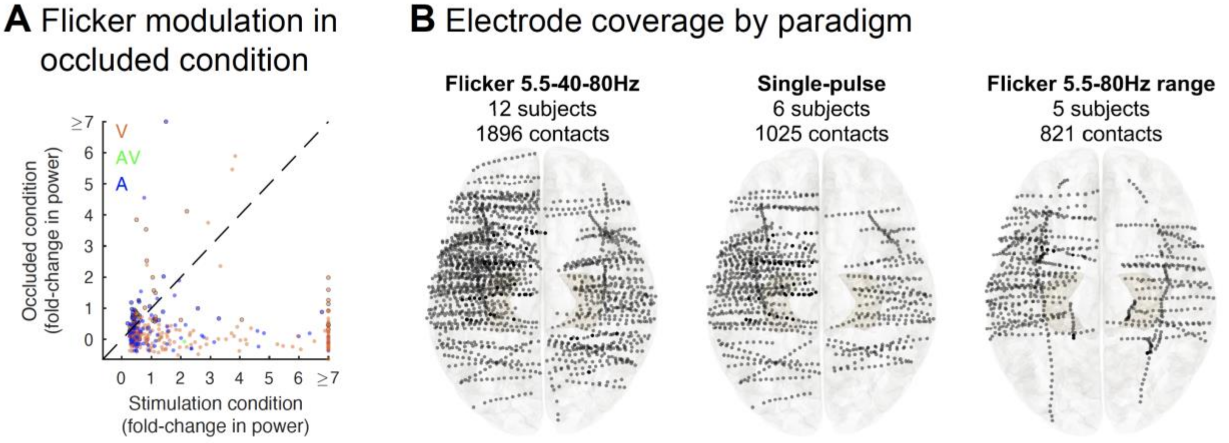


*Figure S1. Relative occluded condition modulation and electrode coverage by paradigm*

*(A) Out of contacts that showed significant flicker modulation to 40Hz visual, auditory or audiovisual flicker in the Flicker 5.5-40-80Hz paradigm, we represented the corresponding fold-change in power (capped at 7) at the frequency of stimulation for the relative occluded condition versus the non-occluded condition. Each dot indicates a contact's responses for a given modality in one recording session with orange, green and blue dots representing visual (V), audio-visual (AV), or auditory (A) stimulus conditions, respectively, and dots circled in black representing results that are significant in the relative occluded condition. Significant modulation in the occluded condition may suggest our occluded condition is not completely successful in occluding sensory stimuli from the subject's visual and auditory systems. For rare cases where we observed a clear peak at the frequency of stimulation in the PSD for the occluded condition, in the majority of those cases the peak was smaller than in the non-occluded condition, which suggests it may be true sensory modulation from imperfect occlusion of the sensory stimuli, rather than noise from the flicker device. Overall, the vast majority of contacts show stronger modulation in the non-occluded condition. This indicates low noise levels using our experimental and preprocessing methods.*

*(B) Number of subjects and number of contacts, as well as approximate location of each contact (represented by dots) across patients on Montreal Neurological Institute (MNI) normalized 3D brain (top view) for each of the three paradigms tested (see Figure 1B, 4B, and 5A for details).*

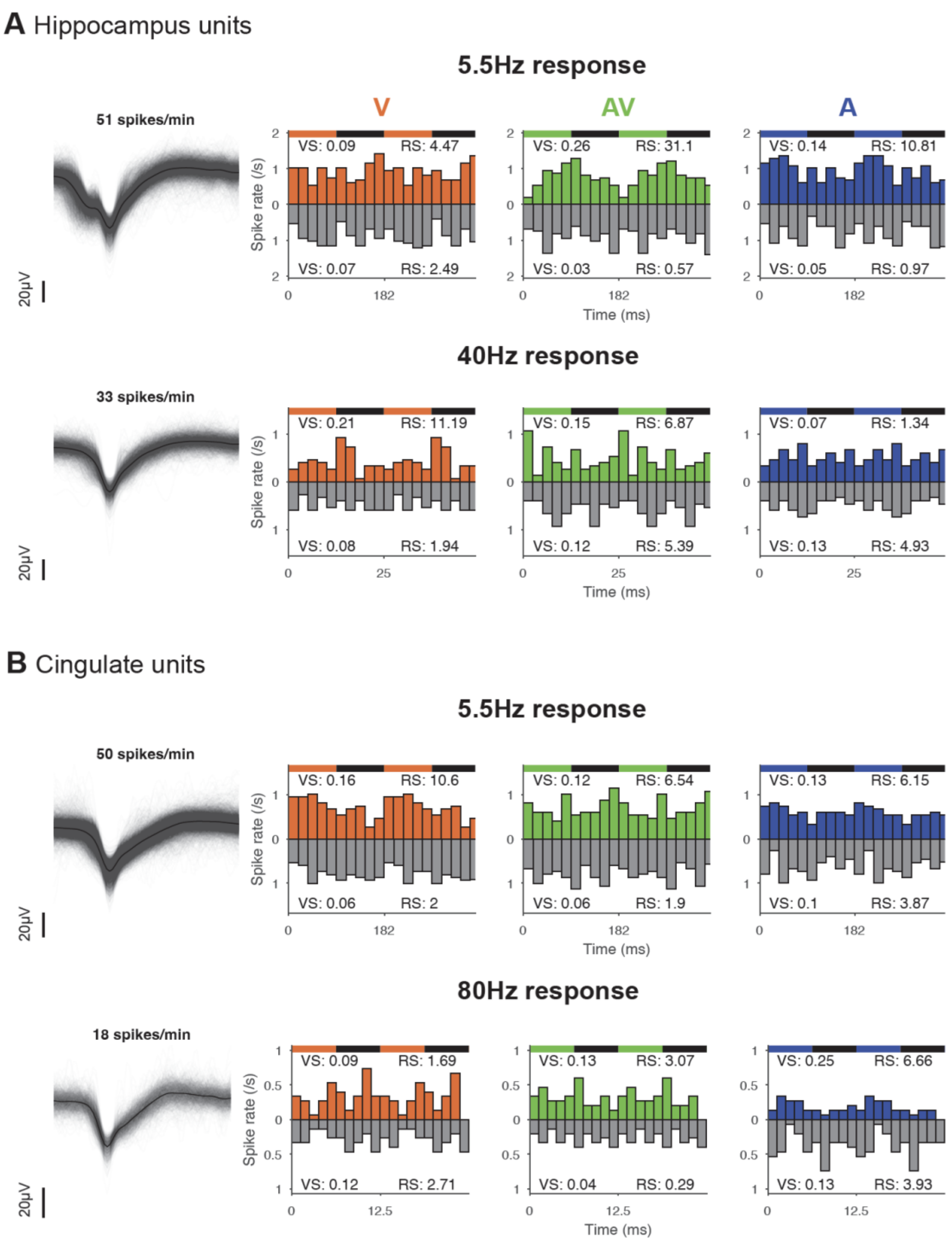


<u>*Figure S2. Flicker modulates neurons' spiking activity in the human hippocampus and cingulate*</u>
*(A) Example single neuron waveforms (left; solid line represents average waveform, transparent lines represent individual waveforms), with peristimulus-time histograms (right) averaged over 2 cycles of the stimulus, illustrating from left to right response to 5.5Hz visual (V, orange), audio-visual (AV, green), and auditory (A, blue) stimulation (colored bars) versus random condition (grey inverted bars). Vector strength (VS) and Rayleigh statistics (RS) for each condition are indicated on the top and bottom of the plot. We see a higher average firing rate at a given phase of the*

*stimulus for the AV condition, showing that this unit is more strongly modulated by 5.5Hz-AV flicker. Bottom: same illustration for a hippocampal multi-unit, in response to 40Hz flicker. This unit seems to be more strongly modulated in the visual modality.*

*(B) Same as (A) for cingulate units, showing response to 5.5Hz flicker (top) and 80Hz flicker (bottom). Top shows single neuron with stronger modulation to 5.5Hz-V stimulation, while bottom shows multi-unit with stronger modulation to 80Hz-A stimulation.*

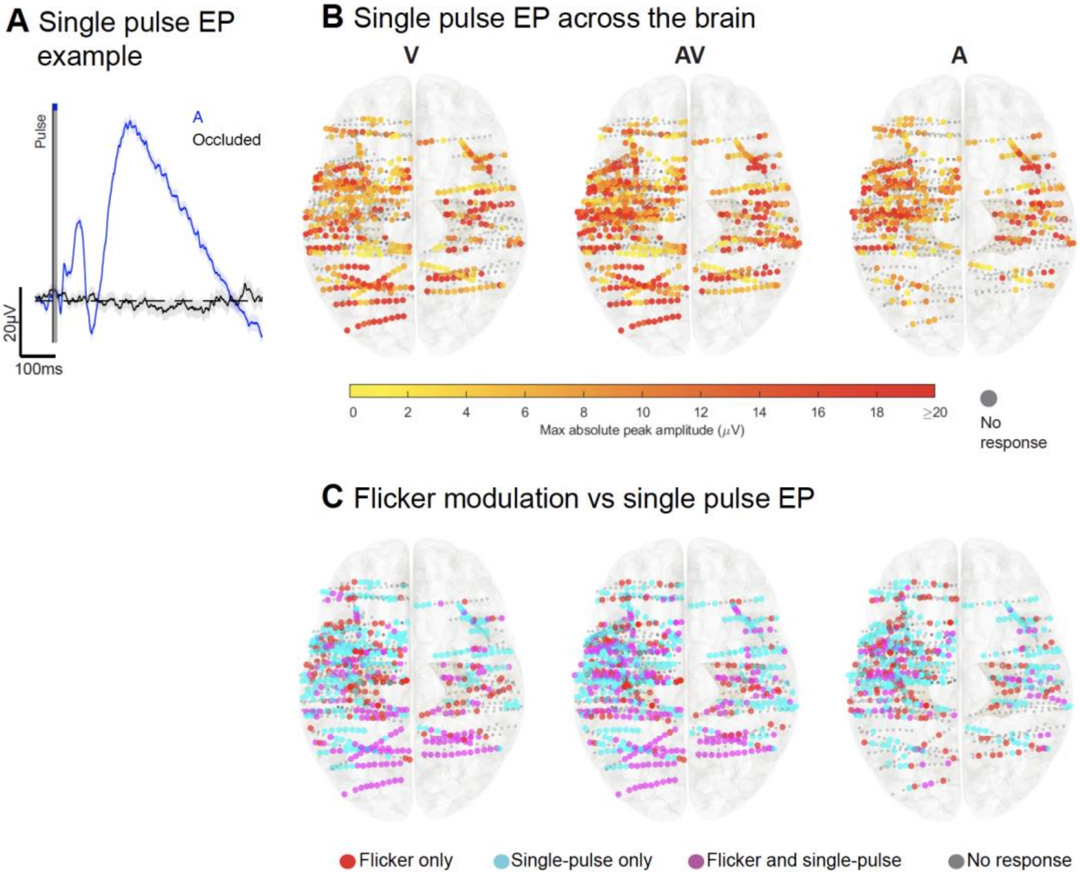


*<u>Figure S3. Single-pulse evoked potential across the brain</u>*

*(A) Example evoked potential (EP), averaged across 200 trials, to auditory (A, blue) versus relative occluded audio-visual (black) pulses, in the primary auditory cortex; solid line represents the mean, shaded area represents standard error of the mean. As expected, we see a rapid (first peak ~20ms), large (up to ~50$\mu$V) response to auditory pulse compared to the relative occluded condition.*

*(B) Approximate location and associated single pulse EP amplitudes of contacts (illustrated with dots) represented on 3D Montreal Neurological Institute (MNI) normalized brain (top view), for visual (V, left), audio-visual (AV, center) and auditory (A, right) modalities, capped at 20$\mu$V.*

*Smaller grey dots represent non-significant single pulse EP responses, while large dots represent significant responses, with maximal absolute peak from 0$\mu$V (yellow) to 20$\mu$V or more (red). There were 97, 148 and 71 contacts with amplitude values higher than 20$\mu$V respectively in the visual, audio-visual, and auditory modalities. As expected, we see a strong response to conditions involving the visual modality in the occipital region, but also in the parietal, temporal, and prefrontal regions. Strong responses to the auditory condition were observed in the temporal region, but also the prefrontal region.*

*(C) Responses to single-pulse versus flicker: approximate location of contacts (represented by dots) and their responses to visual (left), audiovisual (middle) and auditory (right) modalities, represented on 3D MNI brain (top view). Contacts show responses to flicker-only (red), single pulse-only (cyan), both flicker and single pulses (purple), or no response (grey).*

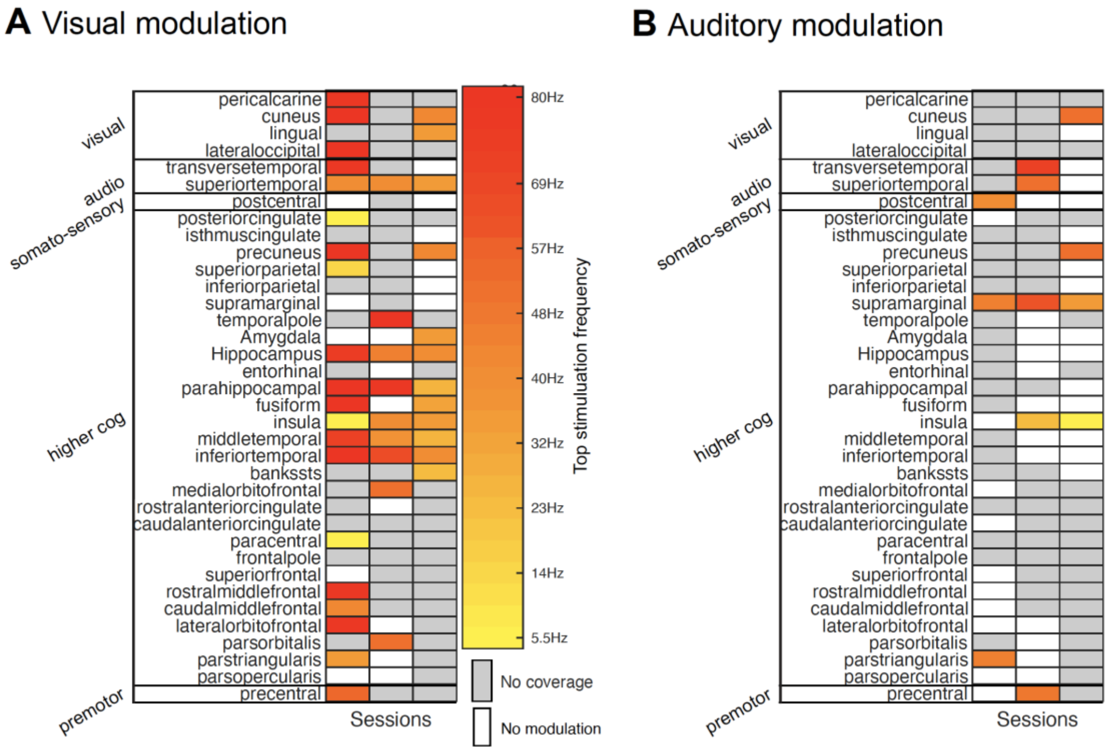


*<u>Figure S4. Preferred stimulation frequency by brain region and modality</u>*

*(A) Representation of the stimulation frequency leading to maximal fold-change in power in each respective brain region, in the case of visual stimulation. Only contacts showing significant fold-change in power to more than six of the stimulation frequencies tested, were included in the analysis. Moreover, when multiple of such channels were located to a given region, the channel responding to the highest number of frequencies, was picked in order to determine top stimulation frequency for that region.*

*(B) Same as (A) but for sessions involving auditory stimulation.*
